# Integrative genetic analysis of the amyotrophic lateral sclerosis spinal cord implicates glial activation and suggests new risk genes

**DOI:** 10.1101/2021.08.31.21262682

**Authors:** Jack Humphrey, Sanan Venkatesh, Rahat Hasan, Jake T. Herb, Katia de Paiva Lopes, Fahri Küçükali, Marta Byrska-Bishop, Uday S. Evani, Giuseppe Narzisi, Delphine Fagegaltier, NYGC ALS Consortium, Kristel Sleegers, Hemali Phatnani, David A. Knowles, Pietro Fratta, Towfique Raj

## Abstract

Amyotrophic lateral sclerosis (ALS) is a progressively fatal neurodegenerative disease affecting motor neurons in the brain and spinal cord. We used 380 post-mortem tissue RNA-seq transcriptomes from 154 ALS cases and 49 control individuals from cervical, thoracic, and lumbar spinal cord segments to investigate the gene expression response to ALS. We observed an increase in microglia and astrocyte expression, accompanied by a decrease in oligodendrocytes. By creating a gene co-expression network in the ALS samples, we identify several activated microglia modules that negatively correlate with retrospective disease duration.

We map molecular quantitative trait loci and find several potential ALS risk loci that may act through gene expression or splicing in the spinal cord and assign putative cell-types for *FNBP1, ACSL5, SH3RF1* and *NFASC*. Finally, we outline how repeat expansions that alter splicing of *C9orf72* are tagged by common variants, and use this to suggest *ATXN3* as a putative risk gene.

## Introduction

Amyotrophic lateral sclerosis (ALS) is a progressively fatal neurodegenerative disease affecting upper and lower motor neurons that control voluntary movement via the corticospinal tract. Most patients have a disease onset in middle age but there is a wide clinical variability in onset of symptoms and the pace of disease progression before death. Disease initiation is thought to occur at a particular point in the spinal cord, brainstem or motor cortex, manifesting with initial clinical symptoms of weakness in a particular limb, or in speech and swallowing. Neurodegeneration then spreads both vertically and laterally within the spinal cord (Ravits and La Spada 2009). The degeneration and death of motor neurons is accompanied by the hallmark TDP-43 protein pathology in 98% of patients (Neumann et al. 2006).

5-10% of ALS cases have a family history of disease (Byrne et al. 2011), with the remaining patients deemed to be sporadic. The field has focused on rare mutations of large effect size, such as large repeat expansions in the gene *C9orf72*, found in not only 40% of familial ALS but also in 10% of sporadic ALS cases (Majounie et al. 2012). Other rare mutations, in genes such as *SOD1, TARDBP, FUS, NEK1, TBK1*, and *KIF5A* make up only a small fraction of the total familial ALS population (Renton, Chiò, and Traynor 2014; Cirulli et al. 2016; Kenna et al. 2016; Nicolas et al. 2018) and the majority of non-familial ALS cases have no known causative mutation. Large-scale genome-wide association studies have repeatedly found common genetic variants associated with ALS risk (van Es et al. 2009; Van Rheenen et al. 2016; Nicolas et al. 2018), making it a complex or polygenic disease. Common polymorphic short-tandem repeats are a further contributor to genetic risk of ALS, the most prominent being *ATXN2*, whereby intermediate lengths impart a small increase in ALS risk (Elden et al. 2010). Since this finding, other members of the *ATXN* gene family have been associated with ALS risk (Tazelaar et al. 2020; Lattante et al. 2018; Hirano et al. 2018). The interplay between rare and common genetic variants in shaping ALS risk is still being explored. Crucially, there has been little progress in assigning risk genes to particular cell-types. One method to achieve this is the mapping of molecular quantitative trait loci (QTLs), the association between common genetic variants and a molecular phenotype such as gene expression. By performing this in a relevant tissue, QTL variants can be colocalized with GWAS risk variants to identify risk genes (Giambartolomei et al. 2014). In Alzheimer’s disease, multiple studies have applied this framework to identify multiple disease risk variants as acting through gene expression and/or splicing in genes specific to microglia and monocytes (Lopes et al., 2021; Young et al. 2021; Novikova et al. 2021).

Although spinal motor neurons are thought to be the primarily affected cell-type in ALS, much research has focused on non-neuronal contributions to disease initiation and progression. Studies using mouse models of *SOD1* mutations have identified a non-neuronal contribution to disease initiation and length of survival (Pramatarova et al. 2001; Jaarsma et al. 2008). These studies and many others identified both astrocytes and microglia as being able to modify disease duration (Yamanaka et al. 2008; Lepore et al. 2008; Boillée et al. 2006; Wang et al. 2009; Phatnani et al. 2013). As motor neurons degenerate during disease they release factors which cause microglia to assume an activated pro-inflammatory state (Town, Nikolic, and Tan 2005; Chiu et al. 2013), which can then induce an activated state in astrocytes (Liddelow et al. 2017). Both activated microglia and astrocytes are toxic to motor neurons (Zhao et al. 2004; Haidet-Phillips et al. 2011), and blocking this microglia-astrocyte crosstalk extends survival in a *SOD1* mouse model (Guttenplan et al. 2020). Several studies have profiled gene expression in human post-mortem ALS tissues, in spinal cord (D’Erchia et al. 2017; Brohawn, O’Brien, and Bennett 2016; Andrés-Benito et al. 2017), frontal cortex (Andrés-Benito et al. 2017), and motor cortex (Dols-Icardo et al. 2020). These studies have identified a broad upregulation of inflammatory and immune-related genes and a downregulation in oligodendrocyte and neuron genes. Further investigation of glial activation and neuron-glia crosstalk in the context of ALS is therefore required. However due to small sample sizes, these studies have been unable to identify more subtle changes in gene expression, nor to compare these changes with clinically variable traits, or to leverage molecular QTLs.

In this study we combined 380 post-mortem tissue RNA-seq transcriptomes from 203 individuals from three different spinal cord segments to investigate the gene expression response to ALS. We identified widespread changes in cellular composition, with increases in astrocyte and microglia markers and glial activation genes, and a decrease in oligodendrocyte expression. We generated gene co-expression networks for the ALS spinal cord, identifying modules of genes that correlate with clinical traits, observing a negative correlation between length of disease duration and modules containing microglia genes.

We then performed QTL mapping and colocalization with ALS GWAS summary statistics. We identified several genetic loci that may alter ALS risk by acting through gene expression and splicing in the spinal cord. Finally, we integrated our differential expression, deconvolution, and co-expression modules to suggest cell types that the colocalized genes may act through.

## Results

### Cellular composition changes in the ALS spinal cord

We aligned and processed post-mortem RNA-seq data from three spinal cord regions (cervical, thoracic, and lumbar) from 154 subjects with ALS and 49 non-neurological controls from the New York Genome Center ALS Consortium, contributed by 8 different medical centres. All samples went through extensive quality control (**Supplementary Fig. 1-3**). Full clinical and demographic details are in **Supplementary Table 1**. Almost no sample had quantifiable expression (TPM > 1) of known motor neuron markers *CHAT, ISL1*, or *MNX1* (**Supplementary Fig. 4**), in line with the low abundance of motor neurons in the bulk spinal cord.

Performing differential expression between all ALS cases and controls in each spinal cord section, controlling for sequencing batch, submitting site, and other technical factors, we found large numbers of differentially expressed genes (DEGs), with the most identified in cervical spinal cord (**Fig. 1a-c**). Correlation of both the test statistics and log_2_ fold change effect sizes between each region found strong concordance in direction (cervical vs lumbar R = 0.87; cervical vs thoracic R = 0.67; lumbar vs thoracic R = 0.58; **Supplementary Fig. 5**). A small number of DEGs were strongly upregulated (log_2_ fold change > 2, equivalent to a 4-fold increase in mean expression) in all three regions, including *CHIT1, GPNMB*, and *LYZ*. Interestingly, these three genes encode proteins secreted by activated microglia. *CHIT1*, encoding the enzyme chitotriosidase, has been shown to be upregulated in the cerebrospinal fluid (CSF) and plasma of ALS patients (Thompson et al. 2018). *GPNMB*, encoding glycoprotein nonmetastatic melanoma B, is upregulated at the protein level in ALS patient CSF and sera (Tanaka et al. 2012) and is expressed by activated microglia (Hüttenrauch et al. 2018). A common genetic variant within an intron of *GPNMB* has been associated with risk of Parkinson’s disease (Murthy et al. 2017; Nalls et al. 2019). *LYZ* encodes human lysozyme, an antibacterial protein secreted by myeloid cells, not previously linked to ALS. A marker of astrocyte activation, *C3 (Liddelow et al. 2017; Guttenplan et al. 2020)*, was also upregulated in all three tissues, albeit with a lower effect size, whereas *MOBP*, an oligodendrocyte marker gene, was downregulated in all three spinal cord sections (**Fig. 1a**).

**Fig. 1.**
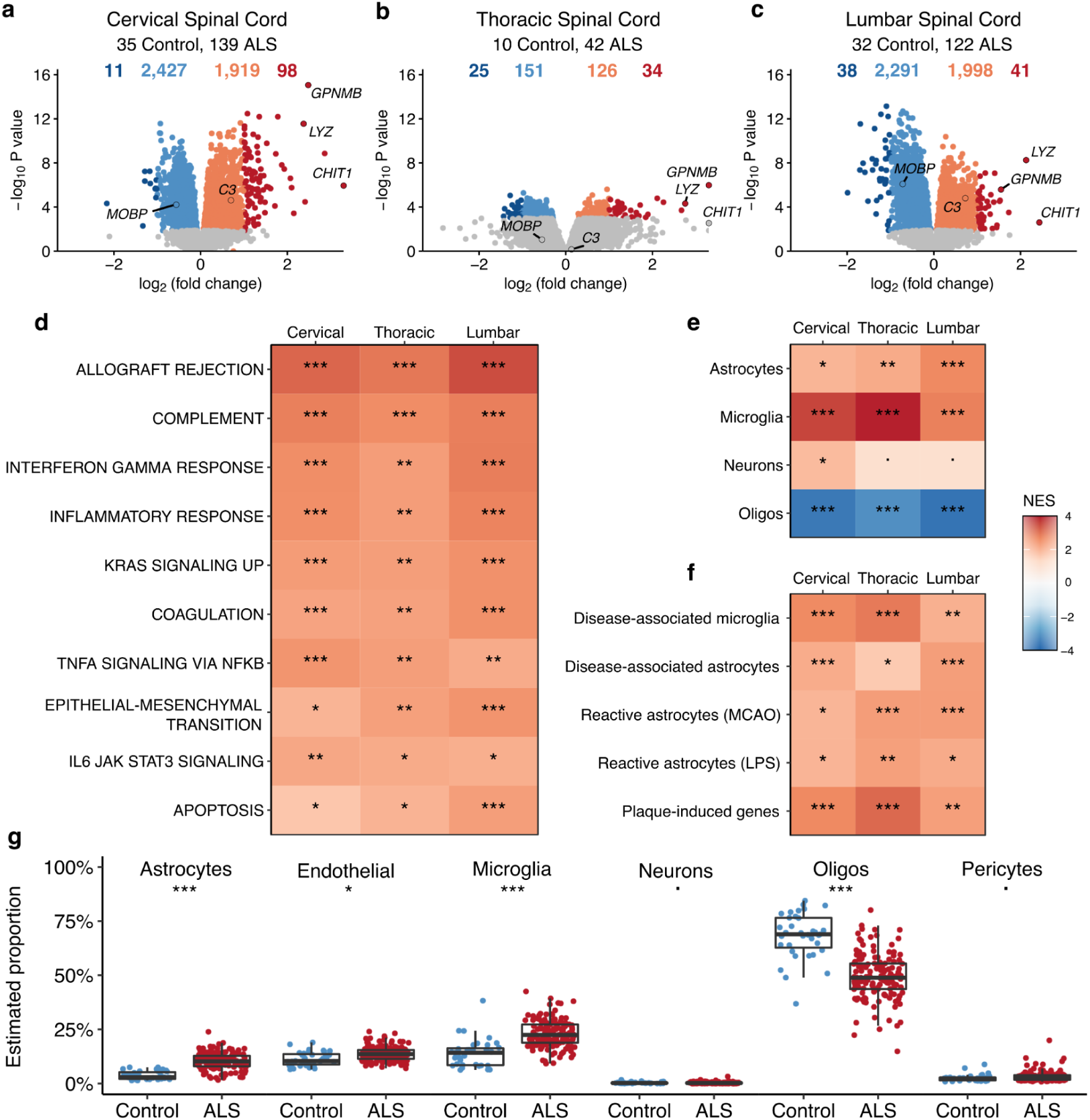
Differential gene expression in the ALS spinal cord is driven by cell-type composition. (**a-c**) Volcano plots comparing ALS patients to controls in each spinal cord section. Genes coloured by whether not differentially expressed (FDR < 0.05; grey), differentially expressed but with modest effects (|log_2_ fold change| < 1; orange) and with stronger effects (|log_2_ fold change| > 1; red). Numbers of genes in each category above the plot. (**d**) GSEA results for the molecular signatures hallmark pathway gene sets. Normalised enrichment score (NES) is a measure of enrichment of a gene set within a ranked list of genes compared to a permuted background. All pathways are enriched in upregulated genes. Significance derived from empirical P-values from a permutation test followed by Bonferroni correction. (**e**) GSEA results for the cell-type signature gene sets. (**f**) GSEA results for the glial activation gene sets. (**g**) Estimated cell-type proportions in the cervical spinal cord, between ALS patients and controls. Significance from a Wilcoxon non-parametric test after regressing technical covariates, followed by Bonferroni correction. ^***^ q < 1e-4; ^**^ q < 1e-3; ^*^ q < 0.05;. q > 0.05. Oligos: oligodendrocytes. Boxplots show the median, first, and third quartiles of the data.

We performed Gene Set Enrichment Analysis (GSEA) (Subramanian et al. 2005) using both curated molecular pathways and sets of cell-type marker genes. Using MSigDB curated pathways (Liberzon et al. 2015), we identified 10 pathways positively enriched in all three tissues (normalised enrichment score (NES) > 1; adjusted P < 0.05), which mostly reflected different immune and inflammatory pathways, with the strongest enrichment observed in allograft rejection, an autoimmune response (**Fig. 1d**). One pathway, cholesterol homeostasis, was negatively enriched in the cervical spinal cord only (**Supplementary Fig. 6)**. We next performed GSEA with lists of the 100 most specific human cell-type marker genes for the four major brain cell-types (Kelley et al. 2018). We observed strong positive enrichment of microglia and astrocyte markers, whilst oligodendrocyte markers were negatively enriched (**Fig. 1e**). There was a modest enrichment in neuronal markers in genes upregulated in the cervical spinal cord, though this was not observed in other tissues.

We then prepared a panel of immune activation genes using four studies of microglia and/or astrocyte responses to pro-inflammatory stimuli in mice. These are disease-associated microglia (Keren-Shaul et al. 2017), disease-associated astrocytes (Habib et al. 2020), activated astrocytes (Zamanian et al. 2012), and plaque-associated genes (Chen et al. 2020). These gene lists only partially overlap (**Supplementary Fig. 7)**, and represent signatures of microglia and astrocyte responses to a range of stimuli, including amyloid plaques, neurodegeneration, hypoxia and lipopolysaccharide. All glial activation sets were enriched in the upregulated genes in all three regions (**Fig. 1f**).

We estimated cell-type proportions in the bulk RNA-seq using both single-nucleus and single-cell RNA-seq from human cortical samples (Mathys et al. 2019; Darmanis et al. 2015), using two different algorithms (Wang et al. 2019; Hunt et al. 2019). The two reference datasets and algorithms produced highly correlated estimates (**Supplementary Fig. 8-11**) and in comparing ALS to controls came to the same broad conclusions as the gene-set enrichment analysis, with the addition of upregulated endothelial cells and pericytes appearing in some but not all of the analyses (**Supplementary Fig. 9**). We present the Mathys estimates for the cervical spinal cord (**Fig. 1g**). As a final analysis of cell-type changes we ran expression-weighted cell-type enrichment (Skene and Grant 2016) using the differentially expressed genes and the same single-nucleus RNA-seq data, which confirmed the observations from deconvolution (**Supplementary Fig. 12**). These results suggest that although any loss of motor neurons is too subtle to be detected in bulk spinal cord samples, there is a robust inflammatory reaction driven by microglia and astrocytes, along with potential dysregulation of oligodendrocytes.

### C9orf72-ALS samples are indistinguishable from sporadic ALS

Analysis of frontal cortex and cerebellum has reported distinct sets of differentially expressed genes between *C9orf72* repeat expansion carriers and sporadic ALS and/or FTD patients (Prudencio et al. 2015; Dickson et al. 2019). We repeated the differential expression analysis but split patients by *C9orf72* repeat expansion status, as assessed by repeat-primed PCR or estimated through ExpansionHunter (Dolzhenko et al., 2019). Comparing each disease set to controls, the directionality of expression changes in each comparison were highly concordant within each spinal cord section (**Supplementary Fig. 13**). Directly comparing *C9orf72* carriers to sporadic ALS cases, no differentially expressed genes were observed, with the exception of the *C9orf72* gene itself, which was downregulated in C9orf72-ALS (cervical spinal cord: log_2_ fold change = -0.45; P = 1e-5). This has been previously observed due to hypermethylation of the *C9orf72* promoter in expansion carriers (Jackson et al. 2020).

### Co-expression network finds disease duration associations

We then created a weighted gene co-expression network using all 303 ALS samples, adjusting for spinal cord region, contributing site and other technical factors. We identified 41 modules (**Fig. 2a; Supplementary Table 5**), and labelled them in ascending order of size from M1 to M41. For each module we created a module eigengene (ME), equivalent to the first principal component of the expression of all genes within that module in each sample (**Supplementary Table 6**). Modules are presented clustered by eigengene correlation (**Fig. 2a**). Co-expression modules are known to identify cell-types (Oldham et al. 2008), and 6 modules were significantly enriched with cell-type marker genes for the major cell types of the brain (**Fig. 2b; Supplementary Table 7**). Using the same panel of glial activation gene sets as before, we found 9 modules enriched for different sets. We observed that the modules enriched with microglia marker genes (M33 and M37) were also enriched for disease-associated microglia and plaque-induced genes, whereas the two astrocyte marker-enriched modules (M18 and M31) were enriched only with disease-associated astrocytes and not with the reactive astrocyte lists. We next performed gene ontology (GO) enrichment on each module. Overall, 39 of 41 modules had at least 1 significant GO term (**Supplementary Table 8**). We manually collapsed GO terms into broad sets (**Fig. 2c**). Some sets reflect probable cell-type specific functions, such as myelination terms with oligodendrocytes, and immune response with microglia, whereas modules enriched in terms relating to gene expression and translation were not enriched with cell-type specific or glial activation markers.

**Fig. 2.**
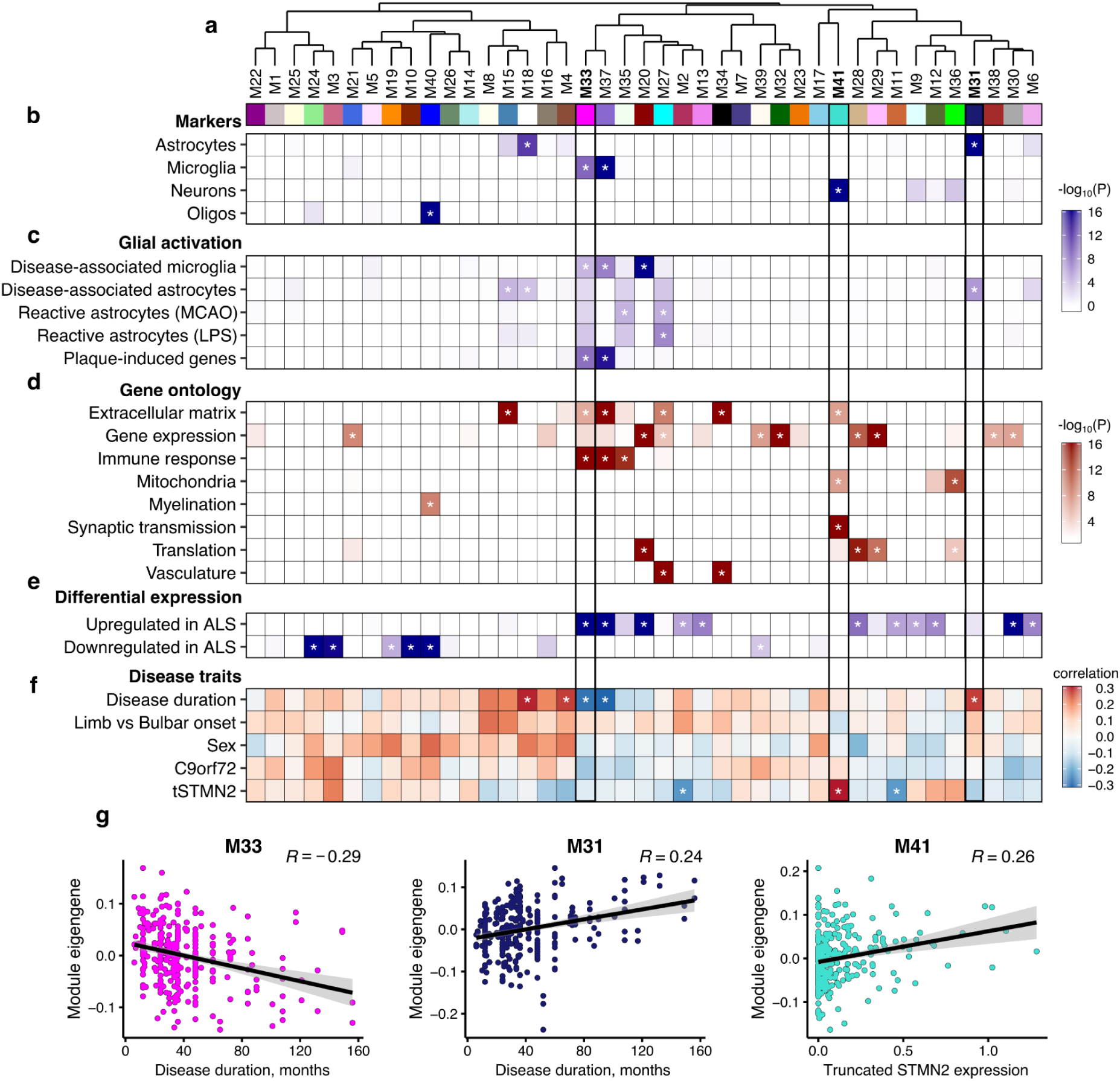
Gene co-expression modules in the ALS spinal cord. **a-g**. Weighted gene co-expression network analysis of 303 ALS spinal cord samples identifies 41 gene modules. **a**. Modules are presented as hierarchical clustering based on module eigengene (ME) correlation. **b-f**. Association results between each module and **b**) cell-type marker genes from Kelley et al, **c**) glial activation genes, **d**) gene ontology (biological process) enrichment, manually collapsed, **e**) differentially expressed genes (FDR < 0.05, no fold change cutoff) between ALS and controls, across all spinal cord regions, **f)** Spearman correlation with disease traits. **g**. MEs for each ALS patient. M33 and M31 correlate with duration of disease in months, and M41 with *tSTMN2* expression. * refers to bonferroni adjusted P < 0.05, per panel. *tSTMN2* - truncated STMN2. TPM - transcripts per million. P-values for b,c,e from one-sided Fisher’s exact test, d from one-sided hypergeometric test, f from Spearman correlation test.

We then used the modules to find associations with clinical variables (**Supplementary Table 9**). Correlating each ME with different clinical traits, we observed 5 modules correlated with retrospective disease duration, defined as the length of time between the age at recorded disease onset and age at death. 2 of the 3 positively correlated modules were enriched with astrocyte marker genes, and both negatively correlated modules were enriched with microglia marker genes. Our previous study using these same samples estimated the abundance of truncated *STMN2* (tSTMN2), a novel cryptic exon transcript created by loss of nuclear TDP-43 (Klim et al. 2019; Melamed et al. 2019). The abundance of *tSTMN2* correlates with levels of phosphorylated TDP-43, and may be a biomarker of TDP-43 pathology (Prudencio et al. 2020). 3 modules correlated with *tSTMN2* abundance. One module, M41, was positively correlated with *tSTMN2* and enriched with neuronal marker genes, including full-length *STMN2*. The two modules negatively correlated with *tSTMN2*, M2 and M11, have no enriched cell-type or ontology but are both enriched for genes upregulated in ALS. No modules were significantly associated with site of onset (limb vs bulbar), sex, or *C9orf72* expansion status.

To further investigate the associations with disease duration, we performed a transcriptome-wide correlation analysis with disease duration as a continuous variable. 650 and 72 genes were significantly associated with disease duration at FDR < 0.05 in the cervical and lumbar spinal cord, respectively (**Supplementary Table 10**). Only 2 significant genes were observed in the thoracic spinal cord, so it was removed from downstream analysis. Test statistics for each gene were highly concordant between the cervical and lumbar cords (Pearson R = 0.71, P < 1e-16; **Supplementary Fig 14**). Using GSEA as before, we found that negatively correlated genes were enriched with microglia markers and microglia activation genes, whereas positively correlated genes were enriched with astrocyte markers but not astrocyte activation gene sets (**Fig. 3b-c**). Using cell-type proportion estimates from the cervical spinal cord, we observed the same negative correlation between duration and microglial proportion (R = -0.31; adjusted P = 0.002), (**Fig. 3d**), but not with astrocyte proportion (R = 0.15; adjusted P = 0.49). *CHIT1* was found to be the strongest negatively correlated gene with disease duration in both cervical and lumbar spinal cord. There is a non-linear relationship between age of onset and age at death in ALS, with shorter durations seen in both younger and older onset patients. We confirm that the association with *CHIT1* expression is strongest with disease duration, and not with age of onset or death (**Fig. 3e-f**).

**Fig. 3.**
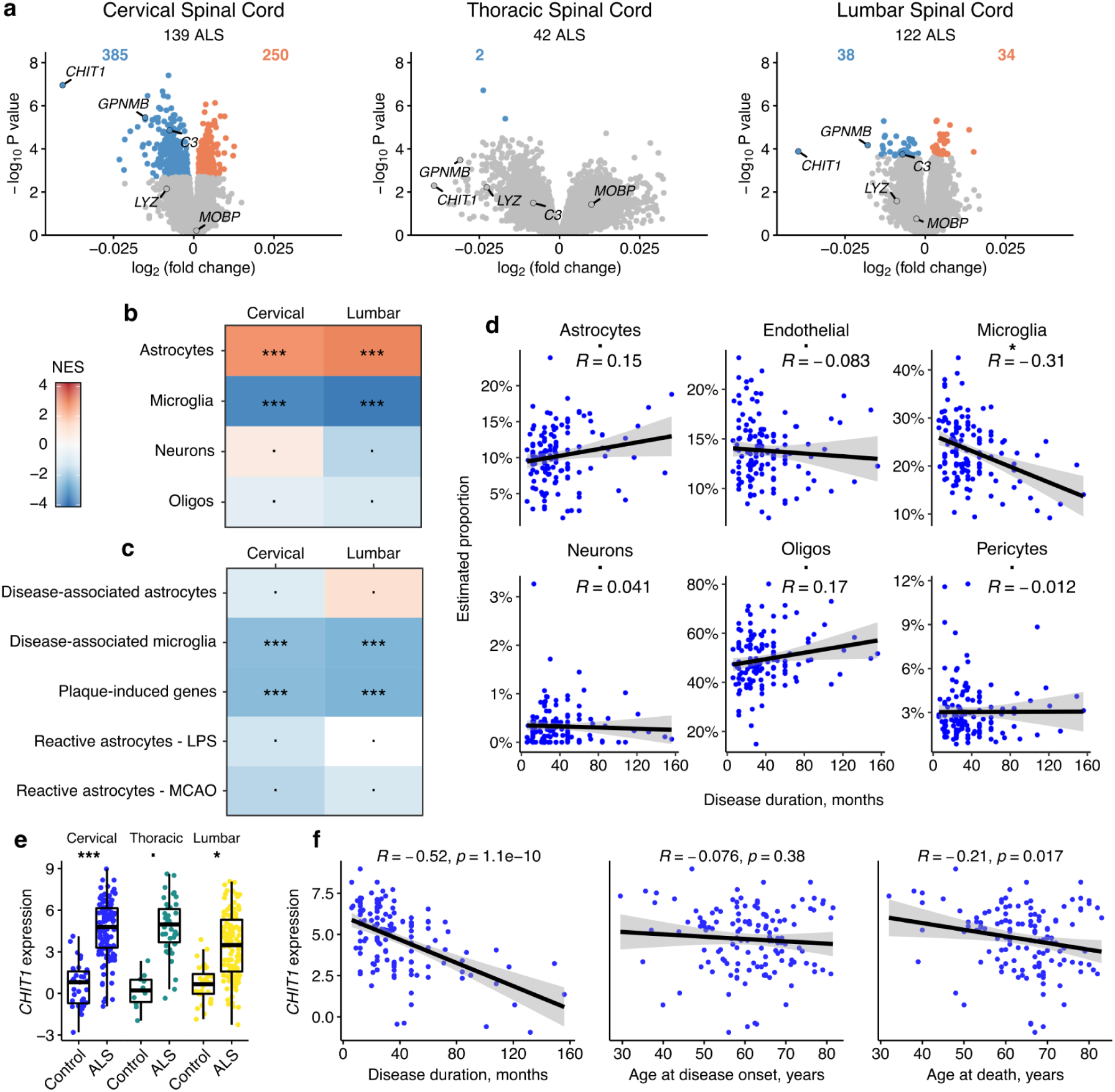
Gene expression correlations with duration of disease. **a**. Volcano plots for correlation in each tissue. **b**. GSEA with cell-type marker genes. **c**. GSEA with glia activation gene lists. **d**. Cell-type proportions in the cervical spinal cord estimated with deconvolution plotted against disease duration. **e**. *CHIT1* is strongly upregulated in ALS in all three tissues. **f**. *CHIT1* expression negatively correlates with disease duration, but not with age of onset, and only weakly with age at death. All correlations are Spearman rank correlations. All P-values in (b-d) are Bonferroni corrected: ^***^ q < 1e-4; ^**^ q < 1e-3; ^*^ q < 0.05;. q > 0.05.

### Mapping spinal cord QTLs

We took common genetic variants (minor allele frequency > 1%) from the matched whole genome sequencing for all donors of European ancestry in the cohort, including cases of non-ALS neurodegeneration. We used this to map quantitative trait loci (QTLs) for gene expression and splicing, the latter using the intron-junction clustering method Leafcutter (Li et al. 2018). We identified 9,492 genes with an expression QTL (eQTL) and 5,627 with a splicing QTL (sQTL) in at least one region (**Fig. 4a**). As a comparison, we downloaded summary statistics for the only other available human spinal cord dataset, from GTEx (v8). We discovered substantially more genes with sQTLs than the 965 found by GTEx. Using Storey’s π_1_ we observed high sharing of QTLs between each region and with GTEx (**Fig. 4b-c**), although sharing was higher in sQTLs than eQTLs, as previously observed (Lopes et al., 2021; The GTEx Consortium 2020). We used our previously generated cell-type proportion estimates to find cell-type interaction QTLs (Kim-Hellmuth et al. 2020) but no tissue had sufficient power to detect any such associations.

**Fig. 4.**
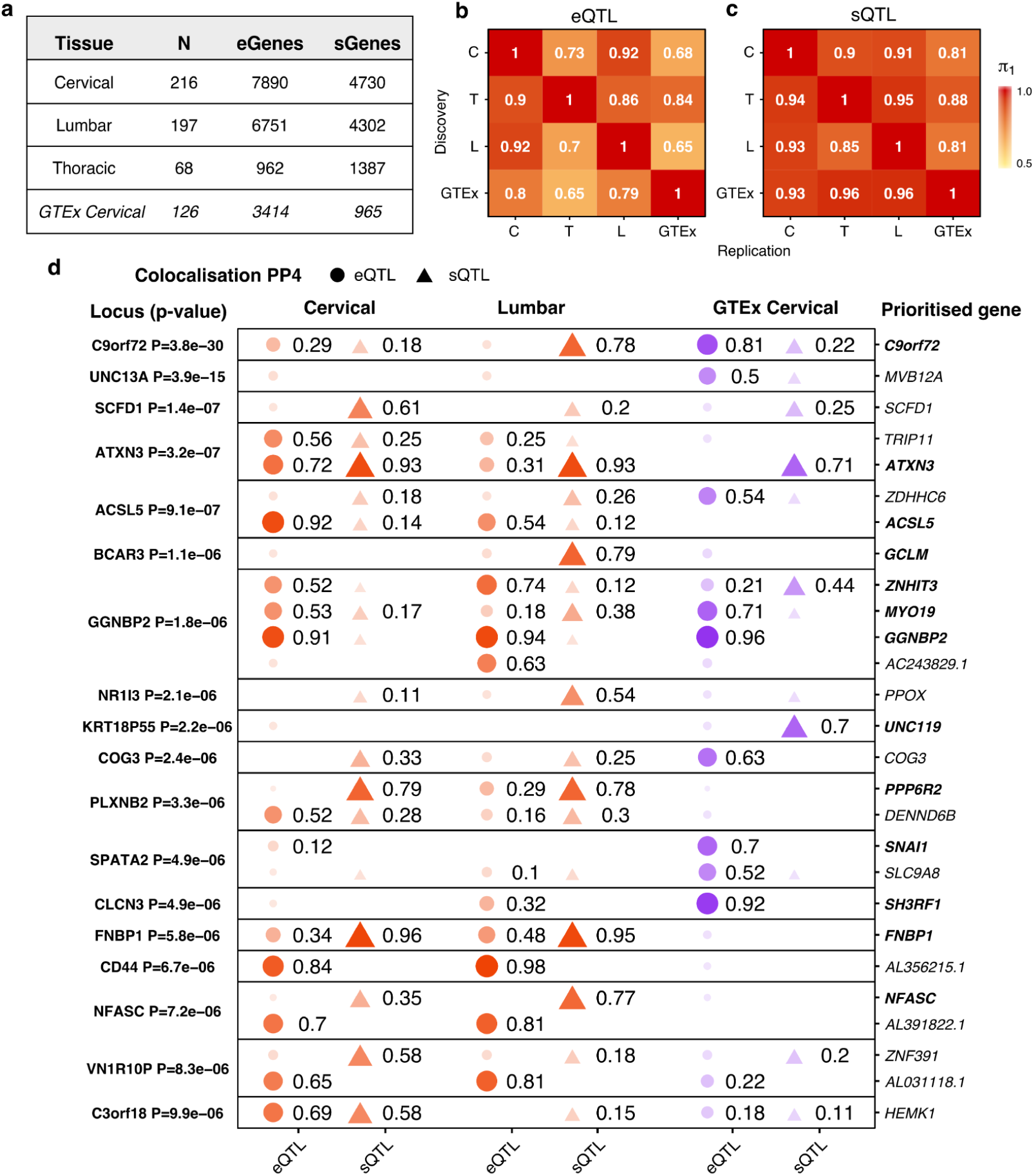
Quantitative trait loci (QTL) colocalize with putative ALS risk variants. **a**. QTL discovery in the three spinal cord tissues and compared with GTEx (v8). Numbers refer to genes with an expression QTL (eGenes) or a splicing QTL (sGenes) at qvalue < 0.05. **b-c**. Sharing of QTLs between tissues using Storey’s π_1_ metric. Values are not symmetric. **d**. Colocalization of subthreshold ALS GWAS loci with spinal cord QTLs. Loci are named for their nearest protein-coding gene. P-values refer to the association of the lead variant in the locus with ALS risk. Numbers refer to the probability of a single shared variant in both GWAS and QTL (PP4). All genes and loci shown with PP4 > 0.5 in at least one QTL dataset. Circles refer to eQTLs, triangles to sQTLs. PP4: posterior probability of colocalization hypothesis 4.

### Putative ALS risk variants colocalise with spinal cord QTLs

We then used our QTLs, in combination with GTEx, to prioritise common genetic risk loci using the latest available ALS GWAS (Nicolas et al. 2018) (**Fig. 4d**). Taking a relaxed approach, we extended our search from the 10 genome-wide significant loci (P < 5e-8) to 64 nominally significant subthreshold loci (P < 1e-5) (**Supplementary Table 12**). Among genome-wide significant loci, we identified strong colocalization with QTLs at a posterior probability of colocalization hypothesis 4 (PP4) > 0.8, only in *C9of72*. In the *UNC13A* locus we observed a potentially spurious colocalization with *MVB12A* only in GTEx. Among the subthreshold loci, we observed colocalization in 16 loci, with the strongest colocalizing genes (PP4 > 0.8) across our tissues and GTEx seen for *ATXN3, GGNBP2, ACSL5* and *FNBP1* (**Supplementary Table 13**).

We then ran transcriptome-wide association study (TWAS), an orthogonal method that uses common variants, gene expression, and splicing ratios to predict cis-regulated expression and splicing. TWAS then imputes those models to GWAS summary statistics to identify genes that are associated with disease risk. We generated TWAS models for each spinal cord section and used summary statistics from the latest available ALS GWAS (Nicolas et al. 2018). In both cervical and lumbar spinal cord, splicing in *C9orf72* and *ATXN3* were significantly associated with ALS (FDR < 0.05) (**Supplementary Fig. 17; Supplementary Table 14**). The lumbar spinal cord TWAS models also identified an association with expression of *MAPT-AS1*, and splicing of *LINC02210* and *LINC02210-CRHR1*. These three genes are within the contentious MAPT H1/H2 haplotype region, which has a complex linkage disequilibrium structure, and so are potential false positives. As a comparison, we downloaded pre-computed expression and splicing weights for the dorsolateral frontal cortex (n = 453; (Li et al. 2019)), which found associations with *C9orf72* in both splicing and expression. In addition, the cortex TWAS models identified *SLC9A8, G2E3, SCFD1*, and *GPX3 (***Supplementary Fig. 17**).

### Annotating prioritised genes to cell-types

We took each colocalised protein-coding gene (PP4 > 0.7) in any of the three spinal cord datasets and looked to annotate cell-type, and to understand how these genes might be involved in ALS. We first took cell-type fidelity ratings from Kelley et al, expressed as a fidelity score from 0-100, with high scores suggesting greater cell-type specificity. Although most genes showed no preference towards any cell type, *FNBP1* (fidelity = 92) showed high specificity to oligodendrocytes (**Fig. 5a**; **Supplementary Fig. 19**). We then used the ALS co-expression network modules generated earlier to infer roles for the genes specifically in ALS. Using guilt-by-association, if a gene belongs to a module enriched in a particular cell-type or marker list, it may also be involved in that cell-type. Both *FNBP1* and *SH3RF1* were placed in module M40, highly enriched for oligodendrocytes (**Fig. 5b**). *NFASC* was placed within M18, a module enriched in both astrocyte marker genes and in disease-associated astrocytes, whereas *ACSL5* was located in M20, a module enriched in disease-associated microglia genes but not microglia markers. We then correlated each prioritised gene with estimated cell-type proportions for six cortical cell types (Mathys et al. 2019). A positive correlation with a particular cell-type proportion is suggestive evidence for specificity. *FNBP1, SH3RF1*, and *NFASC* all positively correlated with oligodendrocyte proportions (**Fig. 5c**). *ACSL5* positively correlated with microglia, endothelial and pericyte proportions, with the strongest correlation seen with endothelial cells. Repeating the analysis in just the control samples replicated the correlations between *FNBP1* and oligodendrocytes and *ACSL5* with endothelial cells (**Supplementary Fig. 20**).

**Fig. 5.**
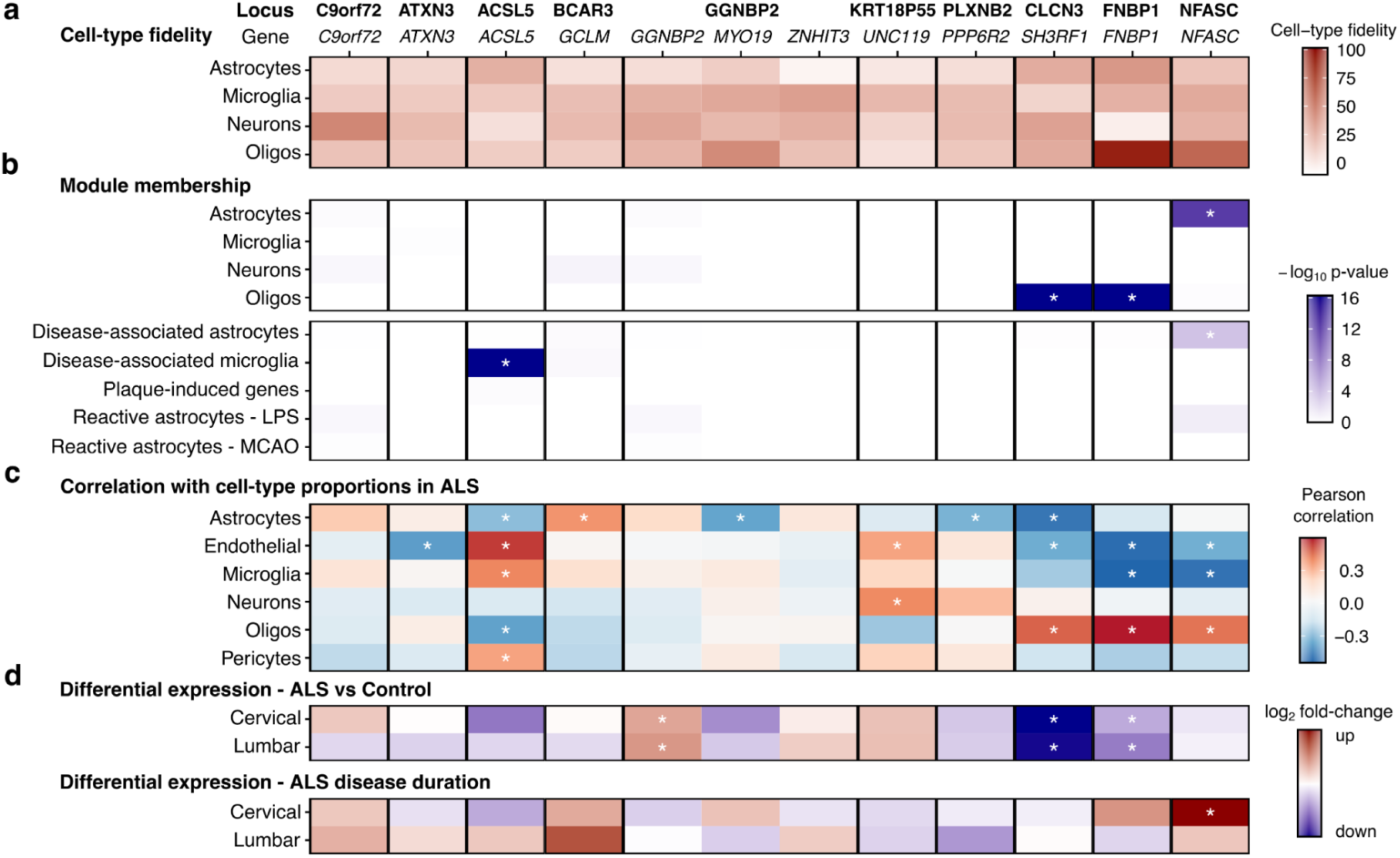
Annotating colocalised genes with cell-type information. **a-d**. Each protein-coding gene with PP4 > 0.7 in at least one spinal cord QTL dataset. **a**. Cell-type fidelity scores from Kelley et al., higher scores imply higher cell-type specificity. **b**. The cell-type and activation marker enrichment p-values from Fig. 4 for the modules containing each gene. **c**. Each gene correlated with estimated cell-type proportions in cervical spinal cord in the ALS samples only. **d**. Log_2_ fold changes from differential expression in ALS vs Control (upper panel) and ALS disease duration (lower panel) in cervical and lumbar spinal cord. ^*^ Bonferroni-adjusted p-value < 0.05.

Finally, looking at the differential gene expression between ALS and Controls, both *FNBP1* and *SH3RF1* are downregulated in ALS cases, whereas *NFASC* expression is positively associated with disease duration in the cervical spinal cord, the only colocalised gene to do so (**Fig. 5d**). *GGNBP2* was upregulated in ALS patients but did not show a clear cell-type specificity. Despite *C9orf72* being highly expressed in mouse microglia (O’Rourke et al. 2016), we observed no associations between *C9orf72* and any cell-type or module.

### Splicing QTLs implicate repeat expansions in ALS risk

The *C9orf72* gene produces transcripts from two alternative promoters, exon 1a and exon 1b. The ALS-associated G_4_C_2_ hexanucleotide repeat expansion (HRE) is located between the two exons (**Fig. 6a**), with more than 30 copies of the HRE considered to be pathogenic (Renton et al. 2011). The *C9orf72* GWAS locus colocalizes with a splicing QTL in the *C9orf72* transcript in the NYGC lumbar spinal cord, as well as an eQTL in GTEx (**Fig. 4a**). The sQTL increases the usage of the intron J1 connecting exon 1a with exon 2, which spans the HRE (**Fig. 6a**). The lead GWAS SNP rs8349943 and the lead sQTL SNP rs1537712 are in strong LD in Europeans (R^2^ = 0.75) and we show that the GWAS SNP is also associated with J1 intron usage (**Fig. 6b**). The lead GWAS SNP rs8349943 is known to tag a founder haplotype which is more susceptible to the HRE (DeJesus-Hernandez et al. 2011). Using ExpansionHunter to estimate the length of the HRE in our cohort, we replicate this finding, as carriers of rs8349943 are also enriched for the HRE (**Fig. 6c**). The usage of the J1 intron is correlated with repeat length (**Fig. 6d**). Therefore, the sQTL colocalization result is likely being driven by the effect of the tagged repeat expansion on the splicing of intron J1.

**Fig. 6.**
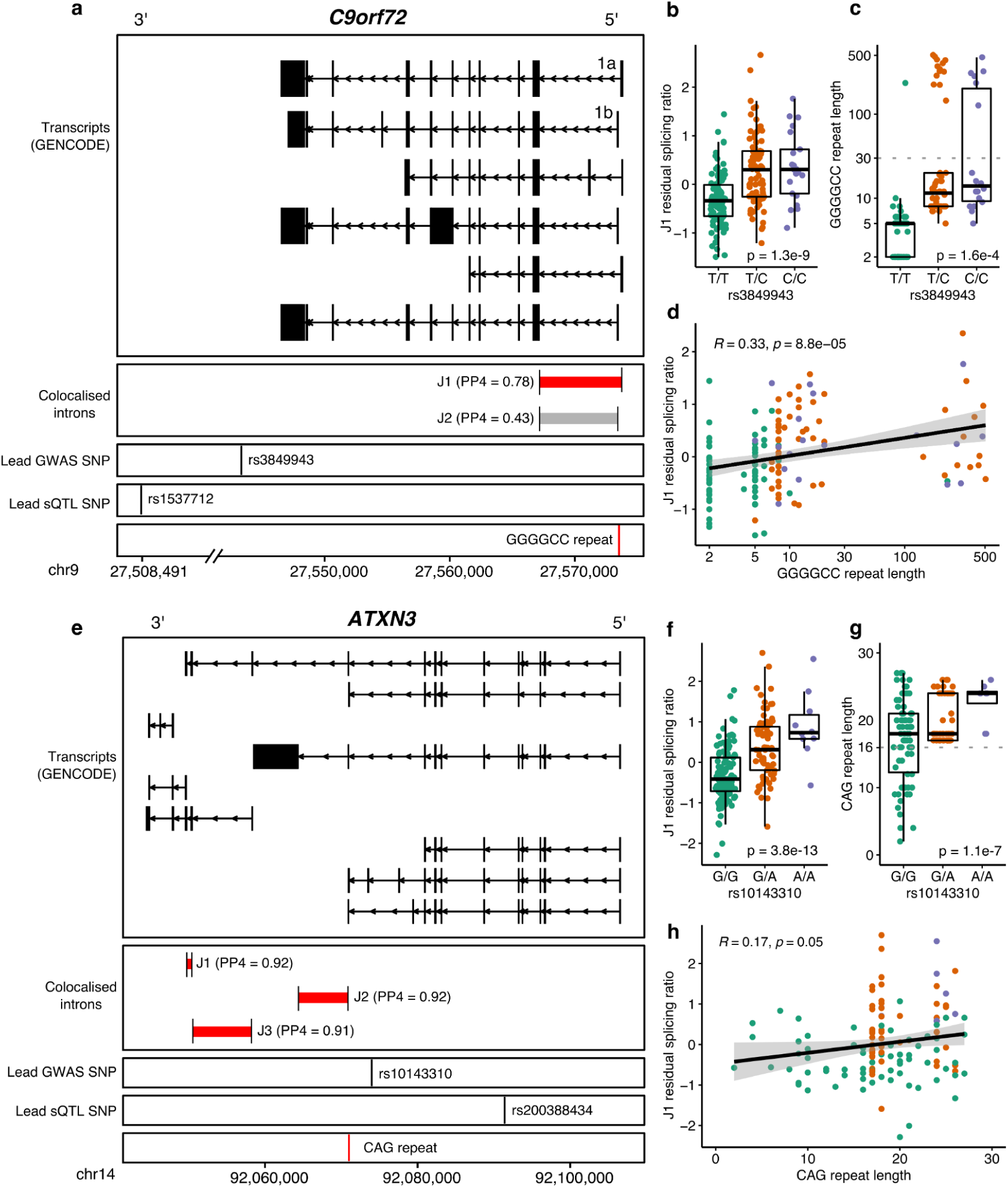
Splicing QTLs illuminate genetic associations with repeat expansions in *C9orf72* and *ATXN3*. **a**. The ALS-causing GGGGCC repeat expansion lies in between the two first exons, 1a and 1b. The intron connecting the exon 1a with exon 2 (J1) has an sQTL in the lumbar spinal cord that colocalises with ALS risk (PP4 = 0.78). **b**. The lead GWAS SNP rs8349943 is associated with J1 intron splicing in the lumbar spinal cord (P = 1.3e-9), linear regression). **c**. The GGGGCC expansion is only observed in carriers of rs8349943. 30 copies of the repeat is considered the threshold for disease initiation (Renton et al. 2011). **d**. The GGGGCC repeat expansion is associated with J1 intron splicing. **e**. The *ATXN3* gene produces multiple transcripts, including several short transcripts at the 3’ end of the gene. Three introns have sQTLs that colocalise with a subthreshold ALS risk GWAS locus with high PP4. The introns are all immediately downstream of a CAG repeat within exon 10. **f**. The lead GWAS SNP rs10143310 is associated with usage of the J1 intron. **g**. Carriers of rs10143310 have a CAG repeat length > 16 copies. **h**. The length of the CAG repeat correlates with J1 splicing (p = 0.05, Spearman correlation).

We propose a similar mechanism for the colocalization of a subthreshold GWAS locus (P = 3.2e-7) with the splicing of *ATXN3*, a promising potential ALS risk gene. The lead SNP rs10143310 fell just short of genome-wide significance in the European ALS GWAS (Nicolas et al. 2018) but did cross the threshold in a multi-ethnic meta-analysis (Nakamura et al. 2020). A CAG repeat in exon 10 of *ATXN3* is highly polymorphic, and expansions greater than 45 copies cause spinocerebellar ataxia type 3 (SCA3), also known as Machado-Josephs disease (Paulson 2006). SCA3 patients have lower motor neuron loss and have detectable TDP-43 protein inclusions (Seidel et al. 2010). Intermediate length expansions, not sufficient to cause ataxia, have been shown to increase ALS risk in several other ataxin family genes, most notably *ATXN2* (Elden et al. 2010), *but also ATXN1* (Tazelaar et al. 2020) *and ATXN8OS* (Hirano et al. 2018). *The idea of tagging repeat expansions in ATXN3* with common genetic variants has been previously explored in the context of SCA3 (Prudencio et al. 2020). In both lumbar and cervical spinal cord, as well as in GTEx, the lead QTL SNP rs200388434 is associated with splicing with a cluster of introns at the 3’ end of the *ATXN3* gene, just downstream of the site of the repeat expansion in exon 10 (**Fig. 5e**). The lead QTL SNP rs200388434 is in high linkage disequilibrium (R^2^ = 0.93) with the lead GWAS SNP rs10143310, and rs10143310 also associated with intron splicing (**Fig. 5f**). Following the example of *C9orf72*, we hypothesised that the GWAS association is tagging an intermediate length CAG repeat, and this may be the underlying causal genetic factor. We were able to genotype the CAG repeat in 304 individuals in the cohort using ExpansionHunter. We observed that the lead QTL SNP is associated with a narrow range of repeat lengths >= 16 (**Fig. 5g**). CAG repeat length also correlated with splicing in the lumbar spinal cord (**Fig. 5h**).

The full NYGC ALS Consortium whole genome sequencing cohort has ExpansionHunter-derived *ATXN3* repeat lengths for 991 ALS cases and 202 controls. When comparing frequencies of each *ATXN3* repeat length we could not observe a significant difference (**Supplementary Fig. 18**), nor by comparing the proportions of samples 21 or more repeats, an arbitrary threshold (31.6% of ALS, 26.2% of controls, P = 0.15).

## Discussion

In this study we assembled the largest ever cohort of post-mortem ALS spinal cords, far surpassing previous studies. This has allowed us not only to identify greater numbers of differentially expressed genes when compared to controls, but to identify genes associated with clinical characteristics within the ALS patient cohort. By integrating common genetic variants we prioritise several new candidate ALS genes that may have cell-type-specific functions. In this way, we can investigate both the cause (genetic risk) and likely consequence (post-mortem gene expression changes) of disease.

Comparing ALS cases to controls we identified robust shifts in cell-type in the three spinal cord regions, primarily composed of a downregulation of oligodendrocytes, and an upregulation in astrocytes and microglia, as well as smaller upward shifts in endothelial cells and pericytes. These observations are seen across the three spinal cord regions and are supported by multiple orthogonal techniques (GSEA, deconvolution, EWCE). However, the interpretation of our results is limited by the use of bulk tissue sections. The reduction in oligodendrocyte gene expression may reflect genuine cell loss due to secondary demyelination accompanying axonal loss (Kang et al. 2013), but this may also reflect a relative shift in proportion compared to increased astrocytes and microglia. For both microglia and astrocytes, although we saw overall upregulation of multiple microglia and astrocyte activation gene lists, it is currently intractable to separate changes in cell-type proportion from changes in cell state in bulk tissue RNA-seq. Therefore the changes we observe could be explained by increased glial proliferation, although the strong effect sizes seen in *LYZ, GPNMB* and *CHIT1* suggest a genuine state shift towards microglia activation. We also cannot rule out that the increased microglia and activated microglia gene expression signatures we observe are not at least partly coming from peripheral monocytes and/or T-cells, which are known to migrate into the spinal cord (Zondler et al. 2016). We also observed small increases in endothelial cells and pericytes. Alterations to the choroid plexus, including reductions in pericytes, have been observed in ALS (Saul et al. 2020). Increases in the recently identified perivascular fibroblast cell-type have been observed in ALS spinal cord RNA-seq as well as ALS mouse models (Månberg et al. 2021), although we did not explicitly look for this cell type.

New approaches will be needed to disentangle effects of tissue composition from cell-intrinsic changes in state, such as single nucleus RNA-seq (Lake et al. 2016) or spatial transcriptomics (Maniatis et al. 2019). The relative paucity of motor neurons in the spinal cord is an obstacle to understanding cell-intrinsic effects in motor neurons, so viable strategies include enriching spinal cord samples for motor neurons (Blum et al. 2021) or for inducing pluripotent stem cells from ALS patients into motor neurons (Ho et al. 2021).

Contrary to a previous study which examined the frontal cortex and cerebellum (Prudencio et al. 2015) we observed no differences between C9orf72-ALS and sporadic ALS spinal cords, with the exception of the *C9orf72* gene. This result is probably affected by the low proportion of motor neurons in the samples, thus masking any motor neuron-specific effects of the expansion. However, as haploinsufficiency caused by the C9orf72 expansion has been shown to affect microglia and macrophages (O’Rourke et al. 2016) we might have expected to see more changes.

Using co-expression networks built in ALS samples only, we observed a series of associations with disease duration and co-expression modules enriched in microglia and astrocyte genes, in opposing directions. Increased numbers of activated microglia, as measured by CD68 staining in the spinal cord, have been observed in faster progressing ALS patients (Brettschneider et al. 2012). However, it is unclear whether microglia activation accelerates neuronal death, or whether microglia activation is an attempted compensatory process, with disease duration driven by some other factor. The negative correlation between *CHIT1* expression and disease duration replicates previous findings at the protein level. Chitotriosidase protein expression is increased in ALS patient CSF, and this higher expression is associated with faster disease progression (Thompson et al. 2018; Varghese et al. 2020). Higher levels of chitotriosidase enzyme activity in blood have been seen in faster progressing ALS patients (Pagliardini et al. 2015). There are further questions about the role of astrocytes. Although M31, a module highly enriched in astrocyte and disease-associated astrocyte genes, positively correlated with disease duration, this was not borne out by deconvolution, suggesting a potential subtype effect. One possible mechanism is that astrocytes may act to stabilise degenerating neurons, increasing survival.

By mapping QTLs we provide a genetic resource for the ALS and wider neuroscience community to understand common genetic drivers of gene expression and splicing in the spinal cord. The unique composition of the spinal cord means QTLs found here could provide insights that cortex-derived datasets lack, especially for diseases driven by glia. One drawback of our study was that we used only donors of European ancestry, and treated each spinal section as an independent dataset. Both of these concessions limited our discovery power. New mixed modelling methods that take account for shared donors and a mixture of ancestries should be able to improve on this (Zeng et al. 2021). We also hope that future studies will be able to combine the NYGC spinal cord samples with GTEx.

Colocalization has allowed us to prioritise new ALS risk genes, but we must stress that the bulk of our findings rely on nominally significant genetic loci. We are also mindful of the potential for false positive associations due to gene co-expression and LD contamination, which affect both colocalization and TWAS (Wainberg et al. 2019). We note that with the recent publication of a new larger ALS GWAS while this manuscript was in preparation (van Rheenen et al. 2021), several of the loci where we prioritised genes by colocalization are now genome-wide significant (*SCFD1, COG3, SLC9A8*). However, the loci we focus on in this study are still below genome-wide significance. We believe that using additional datasets, such as our spinal cord QTLs and TWAS, can overcome the relatively small sample sizes found in ALS GWAS by giving more confidence and credibility to subthreshold GWAS loci.

Our cell-type prioritisation analyses suggest *FNBP1, NFASC* and *SHR3RF1* act primarily within oligodendrocytes. Formin binding protein 1 (*FNBP1)* is involved in formin-mediated actin polymerization (Aspenström 2010), a function shared by Profilin 1 (*PFN1*), a known familial ALS gene (Wu et al. 2012). *FNBP1* reached genome-wide significance in a trans-ethnic meta-analysis of European, Chinese and Japanese ALS GWAS (Nakamura et al. 2020). Neurofascin (*NFASC*) is a immunoglobulin cell adhesion protein that expresses different isoforms in neurons and oligodendrocytes to guide myelination (Nelson and Jenkins 2017). Rare biallelic mutations in *NFASC* cause a neurodevelopment phenotype with both central and peripheral demyelination (Efthymiou et al. 2019). However, its placement within a co-expression module enriched in astrocyte and disease-associated astrocyte genes would suggest it may have other roles. SH3 domain containing ring finger 1 (*SH3RF1)* is a Jun N-terminal kinase (JNK) scaffold involved in neuronal apoptosis. It has been shown to accumulate in models of *CHMP2B*-mediated frontotemporal dementia (West et al. 2018). A recent study on polygenic risk scores in ALS suggested a genetic contribution by oligodendrocytes, which may be mediated by these three loci (Saez-Atienzar et al. 2020).

Acyl-CoA synthetase long chain family member 5 (*ACSL5)* was found within a module enriched in activated microglia and its expression in the ALS spinal cord correlates with endothelial cells and pericytes, as well as microglia. *ACSL5* was previously implicated in astrocyte activation in a canine demyelinating disease (Klemens et al. 2019). The ACSL5 locus was also identified in a trans-ethnic GWAS of European and East Asian individuals (Nakamura et al. 2020). Expression QTLs for *ACSL5* in blood have been associated with ALS risk by Mendelian randomisation (Saez-Atienzar et al. 2020).

By combining our splicing QTLs in *ATXN3* with estimates of the SCA3 expansion, we suggest intermediate repeats in *ATXN3* may increase ALS risk, as has been shown for other ATXN family genes. We were unable to demonstrate this in the whole genome sequencing cohort, but we note that the most recent ataxin repeat gene to be associated with ALS, *ATXN1*, required a large meta-analysis of 7,066 ALS patients and 4,634 controls (Tazelaar et al. 2020). We are therefore underpowered to identify what is likely a small risk modifying effect.

Taken together, our analyses of the ALS spinal cord point to non-neuronal cells as firmly in the heart of the response to disease in the spinal cord, responding to and potentially driving progression of the disease. Our genetic analyses highlight potential new genes that may act on ALS through specific glial cell types. Future genome-wide survival studies may highlight more glial genes in also driving ALS progression. We hope our data are a useful resource for the design of future experiments.

## Methods

### NYGC ALS Consortium cohort

The 1,917 RNA-seq samples from the January 2020 freeze of the New York Genome Center (NYGC) ALS Consortium were downloaded, comprising samples from cortical regions, cerebellum and spinal cord. This study used only spinal cord samples. Diagnosis was determined by each contributing site. Donors include non-neurological disease controls (hereafter controls), those with classical ALS (hereafter ALS), frontotemporal dementia (FTD), mixed pathologies (ALS-FTD, ALS-Alzheimer’s), and a small number of other diseases including Primary Lateral Sclerosis, Kennedy’s Disease and Parkinson’s Disease. *C9orf72* and *ATXN3* repeat expansion lengths were estimated by the Consortium using ExpansionHunter (Dolzhenko et al., 2019) on samples that had PCR-free whole genome sequencing available. Patients with greater than 30 repeats were defined as *C9orf72*-ALS. For ALS patients, age of symptom onset and age at death was reported by each contributing site. Disease duration was defined as the difference between age at death and symptom onset, in months. The NYGC ALS Consortium samples presented in this work were acquired through various institutional review board (IRB) protocols from member sites and the Target ALS postmortem tissue core and transferred to the NYGC in accordance with all applicable foreign, domestic, federal, state, and local laws and regulations for processing, sequencing, and analysis. The Biomedical Research Alliance of New York (BRANY) IRB serves as the central ethics oversight body for NYGC ALS Consortium. Ethical approval was given and is effective through 08/22/2022.

### RNA-seq processing and quality control

The Consortium’s RNA-seq sample processing has been, in part, previously described (Tam et al. 2019; Prudencio et al. 2020). In brief, RNA was extracted from flash-frozen postmortem tissue using TRIzol (Thermo Fisher Scientific) chloroform, followed by column purification (RNeasy Minikit, QIAGEN). RNA integrity number (RIN) (Schroeder et al. 2006) was assessed on a Bioanalyzer (Agilent Technologies). RNA-Seq libraries were prepared from 500 ng total RNA using the KAPA Stranded RNA-Seq Kit with RiboErase (KAPA Biosystems) for rRNA depletion and Illumina-compatible indexes (NEXTflex RNA-Seq Barcodes, NOVA-512915, PerkinElmer, and IDT for Illumina TruSeq UD Indexes, 20022370). Pooled libraries (average insert size: 375 bp) passing the quality criteria were sequenced either on an Illumina HiSeq 2500 (125 bp paired end) or an Illumina NovaSeq (100 bp paired-end). Samples were subjected to extensive sequencing and RNA-Seq quality control metrics at the NYGC that are described below. Notably, a set of more than 250 markers was used to confirm tissue, neuroanatomical regions, and sex in the RNA-Seq data. Only samples passing these metrics are available for distribution. The samples had a median sequencing depth of 42 million read pairs, with a range between 16 and 167 million read pairs.

Samples were uniformly processed using RAPiD-nf, an efficient RNA-Seq processing pipeline implemented in the NextFlow framework (Di Tommaso et al. 2017). Following adapter trimming with Trimmomatic (version 0.36) (Bolduc, 2016), all samples were aligned to the hg38 build (GRCh38.primary_assembly) of the human reference genome using STAR (2.7.2a) (Dobin et al. 2013), with indexes created from GENCODE, version 30 (Harrow et al. 2012). Gene expression was quantified using RSEM (1.3.1) (B. Li and Dewey 2011). Quality control was performed using SAMtools (H. Li et al. 2009) and Picard, and the results were collated using MultiQC (Ewels et al. 2016).

Aligned RNA-seq samples were subjected to quality control modeled on the criteria of the Genotype Tissue Expression Consortium (Consortium and The GTEx Consortium 2020). Any sample failing 1 of the following sequencing metric thresholds was removed: a unique alignment rate of less than 90%, ribosomal bases of greater than 10%, a mismatch rate of greater than 1%, a duplication rate of greater than 0.5%, intergenic bases of less than 10.5%, and ribosomal bases of greater than 0.1%. For tissue identity, both principal components analysis and UMAP were performed on the TMM-normalised gene expression matrix, followed by k-means clustering. This identified three clusters of samples, grouped by cerebellum, cortex and spinal cord. Samples that clustered with a non-matching tissue type were flagged and tissue identity was re-confirmed using the expression of the cerebellar marker *CBNL1*, the cortical marker *NRGN* and the oligodendrocyte marker *MOBP*. 19 samples were removed for having ambiguous tissue identity. For duplicate samples, where samples of the same tissue from the same donor were sequenced, the sample with the highest RIN was retained, this removed 15 duplicate samples. Sex was confirmed using *XIST* and *UTY* expression. 11 samples with missing sex information were confirmed as males. Due to the large impact of RNA integrity number (RIN) on expression, only samples with RIN >= 5 were included in the differential expression analysis, totalling 380 spinal cord samples from 203 donors. For the QTL analyses (see below), no RIN threshold was applied.

### Covariate selection and modelling for differential expression

The following was run for each tissue separately: Clinical variables (disease status, age at death, sex, contributing site) were combined with sequencing variables (RIN, sequencing preparation method, sequencing platform), technical metrics of the RNA-seq libraries from Picard (% mRNA bases, 3’ bias, etc), and genotype principal components (see below). Using voom-normalised gene expression removing lowly expressed genes (genes must have >1 counts per million in at least 75% of samples), principal components analysis was performed. The top 10 principal components were then associated with each potential confounding variable using a linear model, estimating the variance explained (r^2^) of the confounder on each principal component (**Supplementary Fig. 3a**). Using an orthogonal approach, variancePartition (Hoffman and Schadt 2016) was run on a reduced set of confounding variables, taking only the nominally independent sequencing metrics (**Supplementary Fig. 3b)**.

For performing differential gene expression between ALS and control samples, multiple model designs were fitted to account for differences in sequencing batch and contributing site, both of which are correlated with disease status. To account for potentially non-linear dependence of RIN and age at death, squared terms were included. To account for potential confounding differences due to genetic background, the first 5 genotype principal components (gPCs) from EIGENSOFT (Price et al. 2006) were included. For Cervical and Lumbar spinal cord, the following model was fitted: *expression ∼ disease + sex + library preparation method + contributing site + age + age*^*2*^ *+ RIN + RIN*^*2*^ *+ % mRNA bases + % chimeric reads + % ribosomal bases + % intergenic bases + median 3’ bias + median 5’ bias + % read 1 stranding + % adapter + gPC1 + gPC2 + gPC3 + gPC4 + gPC5*. For the smaller set of Thoracic spinal cord samples, a reduced model was fitted as it maximised the gene-gene correlation of differential expression effect sizes with the other two regions: *expression ∼ disease + sex + RIN + RIN*^*2*^ *+ age + age*^*2*^ *+ library preparation method + gPC1 + gPC2 + gPC3 + gPC4 + gPC5*. Differential gene expression was fitted using limma voom (Law et al. 2014) on TMM- and quantile-normalized (Risso et al. 2011) read counts. P-values were adjusted for multiple testing using FDR correction, with genes were considered differentially expressed at FDR < 0.05. A gene was considered to have a large effect size at |log_2_ fold change| > 1.

For transcriptome-wide correlations with age of onset and disease duration, the same models as before were used in the ALS samples only, with either age of onset (years) or disease duration (years) used as continuous variables. Results for each tissue were correlated by matching the log_2_ fold change for each gene and performing Pearson correlation (**Supplementary Fig. 5)**.

### Gene set enrichment analysis

Sets of genes were collected from multiple sources and compared to the full differential expression results for each tissue using Gene Set Enrichment Analysis (GSEA) (Subramanian et al. 2005), as implemented in the Clusterprofiler R package (Yu et al. 2012). As input we included all tested genes from the differential expression or disease duration analysis for each tissue at nominal (unadjusted) P-value < 0.05, ranked by log_2_ fold change. For each gene set, a running cumulative tally is made of whether genes in a set are present or absent during a walk down the list. The maximal score during the walk is the enrichment score (ES), which reflects the degree of which a gene set is enriched at either the top or bottom of a list. Labels are then randomly permuted to generate an empirical null ES distribution and a P-value is calculated. To aid comparison between sets, each ES is then divided by the mean null ES to create a normalised enrichment score (NES). Hallmark pathway gene sets (h.all.v7.2.symbols.gmt) were downloaded from the molecular signatures database (Liberzon et al. 2015). The top 100 high fidelity human marker genes (Kelley et al. 2018), for astrocytes, microglia, neurons, and oligodendrocytes were downloaded from the accompanying resource website (**see URLs**). Disease-associated Microglia (DAM) signature genes (Keren-Shaul et al. 2017), Disease-associated astrocytes (Habib et al. 2020), Plaque-associated genes (Chen et al. 2020), and LPS and MCAO-activated astrocyte genes (Zamanian et al. 2012) were downloaded from their respective supplementary materials. Mouse genes were lifted over to their human homologues using Homologene (Mancarci and French 2019). Any duplicate gene name, or gene name without a matching Ensembl ID in GENCODE v30 was removed.

### Cell-type deconvolution

Filtered counts and cell-type labels for single nucleus RNA-seq from 80,660 cells from 48 human dorsolateral prefrontal cortex samples (Mathys et al. 2019) were downloaded from Synapse (syn18681734). Only cells from the 14 donors without dementia were kept. Single cell RNA-seq data of 466 cells from 12 donors (Darmanis et al. 2015) was downloaded from Gene Expression Omnibus (GSE67835) using the count matrices and cell-type labels provided. Bulk spinal cord RNA-seq data was voom-normalized before deconvolution was estimated using MuSiC (Wang et al. 2019), a state-of-the-art method which incorporates the variance between multiple donors from single cell/nucleus RNA-seq. Cell-type proportion estimates using the two different reference datasets were highly correlated, although the magnitude of the estimates differed substantially (**Supplementary Fig. 8-9**). In addition, we ran dtangle (Hunt et al. 2019) using the Darmanis reference. Estimates from dtangle and MuSiC were highly correlated (**Supplementary Fig. 10-11**). Estimated proportions of each cell-type were compared between ALS and control using non-parametric Wilcoxon tests after regressing the same technical covariates above. P-values were corrected for multiple testing using the Bonferroni correction. For comparing duration of onset, estimated cell-type proportions were correlated using a Spearman correlation.

### Expression-weighted Cell-type Enrichment

Expression-weighted cell-type enrichment analysis was performed using the EWCE package (Skene and Grant 2016). Cell-type specificity scores for each gene were created using human frontal cortex single-nucleus RNA-seq (Mathys et al. 2019). Cell-type enrichment results were generated using the top 250 upregulated and downregulated genes, ordered by t-statistic, for the differential expression results for each segment. Specificity scores for each set were then compared to the mean of the empirical null distribution from 10,000 random gene sets. Enrichment was expressed as the number of standard deviations from the mean. P-values were Bonferroni corrected for multiple testing. Significance was set at adjusted P < 0.05.

### Gene co-expression Networks

Gene expression from all 303 ALS samples from the three spinal cord regions was combined into a single matrix. Genes annotated as protein-coding by Ensembl were kept, and only then if each gene had at least 1 read count per million in at least 50% of samples, resulting in 14,375 genes. Gene counts were then transformed using Voom and TMM normalization. The following covariates were then regressed out using removeBatchEffect(): contributing site, spinal cord section, RIN, % mRNA bases, % ribosomal bases, % intergenic bases, median 3’ bias, median 5’ bias, % chimeric reads, and genomic PCs 1-5.

Co-expression network analysis was performed using Weighted Gene Correlation Network Analysis following a standard pipeline. Scale-free topology (R^2^ > 0.8) was achieved by applying a soft threshold power of 5 into a signed network model. The adjacency matrices were constructed using the average linkage hierarchical clustering of the topological overlap dissimilarity matrix (1-TOM). Co-expression modules were defined using a dynamic tree cut method with minimum module size of 50 genes and deep split parameter of 4. Modules highly correlated with each other, corresponding to a module eigengene (ME) correlation > 0.75, were merged, resulting in a total of 41 modules. Modules were labelled according to their size.

We calculated the Spearman correlation between each module eigengene and the following clinical variables: disease duration (years), site of disease onset (bulbar or limb), *C9orf72* status, sex, and *tSTMN2* abundance. *tSTMN2* abundance in TPM for the matching samples was extracted from the supplementary data from (Prudencio, Humphrey, et al. 2020).

Cell-type and glial activation genes were tested for enrichment within each module using Fisher’s exact test followed by Bonferroni correction. Gene ontology biological process terms were tested for enrichment using the gProfiler2 package (Reimand et al. 2007). Terms with less than 10 genes were removed before correction for multiple testing. Enriched terms were then manually grouped into sets for presentation. Full module assignments, eigengenes, and enrichment results are shared as **Supplementary Tables 5-9**.

### Whole Genome Sequencing (WGS)

Read alignment to the human reference genome GRCh38, duplicate marking, and Base Quality Score Recalibration (BQSR) were performed as described in the functional equivalence pipeline standard developed for the Centers for Common Disease Genomics project (Regier et al. 2018). All 513 samples were jointly genotyped using GATK HaplotypeCaller 3.5 (Poplin et al, 2017). We used GATK’s VariantRecalibrator to train the Variant Quality Score Recalibration (VQSR) model using “maxGaussians 8” and “maxGaussians 4” parameters for SNVs and INDELs, respectively. We applied the VQSR model to the joint call-set using ApplyRecalibration with truth sensitivity levels of 99.8% for SNVs and 99.0% for INDELs. Variants overlapping the ENCODE hg38 blacklist regions (Amemiya, Kundaje, and Boyle 2019) were removed. Variants were then filtered using the following rules inspired by the GTEx v8 consortium (Aguet et al. 2019; The GTEx Consortium 2020), and a pipeline which used empirical thresholds using discordant variant calls between replicate samples (Adelson et al. 2019). The pipeline was implemented in bcftools (Danecek et al. 2021), vcftools (Danecek et al. 2011) and PLINK (Chang et al. 2015) using the snakemake framework (Köster and Rahmann 2012). SNPs and indels were filtered using the following filters: genotype level read depth (GDP) > 10; Genome quality (GQ) > 20; missingness < 15%; Overall read depth (total read depth) > 5000 (equivalent to ∼10x coverage); 58.75 > mapping quality (MQ) > 61.25; variant quality score log-odds (VQSLOD) > 7.81; inbreeding coefficient > -0.8. Finally, a minor allele frequency cut-off of 1% was applied. In total, 6,711,470 SNPs were used for testing. Numbers of SNPs retained at each filtering step are recorded in **Supplementary Table 11**.

LD-pruned biallelic SNPs were selected from the cohort-level VCF and combined with the 1000 Genomes Project (1kGP) phase 3 call set (1000 Genomes Project Consortium et al. 2015). Ancestry was inferred by projecting samples of interest onto genotype PCs computed based on the 1kGP samples using smartpca v6.0.1 (Price et al. 2006). Non-European or admixed samples were flagged if they were 4 standard deviations from the European super-population in the first 2 PCs, which flagged 65 individuals for exclusion from the analysis.

Samples were excluded if they had > 10% individual missingness, which removed no samples. Relatedness was assessed with KING (Manichaikul et al. 2010). Samples were flagged if they had a relatedness > 0.125. 25 pairs of duplicate individuals submitted by different cohorts and a pair of related individuals, were removed. Samples were checked for ambiguous sex genotype, or a mismatch between the DNA sex and the inferred sex from the RNA-seq. 1 individual was removed for having assigned XO/XY mosaic karyotype. Possible sample swaps were assessed by comparing each genotyped individual to inferred SNPs in the RNA-seq, using MBV from QTLtools (Fort et al. 2017). No sample swaps were observed. Across the three spinal cord regions. 481 samples from 236 unique donors were used for QTL mapping.

### Quantitative Trait Loci mapping

To perform expression QTL (eQTL) mapping, we created a pipeline based on the one created by the GTEX consortium. We completed a separate normalization and filtering method to previous analyses. Gene expression matrices were created from the RSEM output using tximport (Love, Soneson, and Robinson 2017). Matrices were then converted to GCT format, TMM normalized, filtered for lowly expressed genes, removing any gene with less than 0.1 TPM in 20% of samples and at least 6 counts in 20% of samples. Each gene was then inverse-normal transformed across samples. PEER (Stegle et al. 2012) factors were calculated to estimate hidden confounders within our expression data. We created a combined covariate matrix that included the PEER factors and the first 5 genotyping principal component values as input to the analysis. We tested numbers of PEER factors from 0 to 30 and found that between 10 and 30 factors produced the largest number of eGenes in each region (**Supplementary Fig. 16**).

To test for cis-eQTLs, linear regression was performed using the tensorQTL (Taylor-Weiner et al. 2019) *cis_nominal* mode for each SNP-gene pair using a 1 megabase window within the transcription start site (TSS) of a gene. To test for association between gene expression and the top variant in cis we used tensorQTL cis permutation pass per gene with 1000 permutations. To identify eGenes, we performed q-value correction of the permutation P-values for the top association per gene at a threshold of 0.05.

We performed splicing quantitative trait loci (sQTL) analysis using the splice junction read counts generated by regtools (Feng et al. 2018). Junctions were clustered using Leafcutter (Li et al. 2018), specifying for each junction in a cluster a maximum length of 100kb. Following the GTEx pipeline, introns without read counts in at least 50% of samples or with fewer than 10 read counts in at least 10% of samples were removed. Introns with insufficient variability across samples were removed. Filtered counts were then normalized using *prepare_phenotype_table*.*py* from Leafcutter, merged, and converted to BED format, using the coordinates from the middle of the intron cluster. We created a combined covariate matrix that included the PEER factors and the first 5 genotype principal components as input to the analysis. We mapped sQTLs with between 0 and 30 PEER factors as covariates in our QTL model and determined 5 and 15 factors produce the largest number of sGenes (**Supplementary Fig. 16**).

To test for cis sQTLs, linear regression was performed using the tensorQTL nominal pass for each SNP-junction pair using a 100kb window from the center of each intron cluster. To test for association between intronic ratio and the top variant in cis we used tensorQTL permutation pass, grouping junctions by their cluster using --grp option. To identify significant clusters, we performed q-value (Storey 2003) correction using a threshold of 0.05.

We estimated pairwise replication (π1) of eQTLs and sQTLs using the q-value R package. This involves taking the SNP-gene pairs that are significant at q-value < 0.05 in the discovery dataset and extracting the unadjusted P-values for the matched SNP-gene pairs in the replication dataset.

### GTEX Spinal Cord QTL summary statistics

Full summary statistics for the cervical spinal cord expression QTLs (v8) were downloaded from the eQTL catalogue (**see URLs**). The splicing QTLs were downloaded from the Google Cloud portal. Top associations for each gene were downloaded from the GTEx portal.

### Genome-wide association study summary statistics

Full summary statistics for the latest ALS GWAS (Nicolas et al. 2018) were downloaded from the EBI GWAS Catalogue, which have lifted over the variants to the hg38 build. Genome-wide significant loci were taken to be the most significant variants within 1 megabase at a threshold of P < 5e-8. Subthreshold loci were defined at a relaxed threshold of P < 1e-5. Loci were named by their nearest protein-coding gene using SNPnexus (Oscanoa et al. 2020).

### Colocalization analysis

We used coloc (Giambartolomei et al. 2014) to test whether SNPs from different loci in the ALS GWAS colocalized with expression and splicing QTLs from the spinal cord. For each genome-wide and subthreshold locus in the ALS GWAS we extracted the nominal summary statistics of association for all SNPs within 1 megabase either upstream/downstream of the top lead SNP (2Mb-wide region total). In each QTL dataset we then extracted all nominal associations for all SNP-gene pairs within that range and tested for colocalization between the GWAS locus and each gene. To avoid spurious colocalization caused by long range linkage disequilibrium, we restricted our colocalizations to GWAS SNP - eQTL SNP pairs where the distance between their respective top SNPs was ≤ 500kb or the two lead SNPs were in moderate linkage disequilibrium (r^2^ > 0.1), taken from the 1000 Genomes (Phase 3) European populations using the LDLinkR package (Myers, Chanock, and Machiela 2020).

For splicing QTLs we followed the same approach but collapsed junctions to return only the highest PP4 value for each gene in each locus. Due to the smaller window of association (100kb from the center of the intron excision cluster) we restricted reported colocalizations to cases where the GWAS SNP and the top sQTL SNP were either within 100kb of each other or in moderate linkage disequilibrium (r^2^ > 0.1).

### Estimating repeat expansions with ExpansionHunter

ExpansionHunter (v2.5.5) was employed to estimate the length of disease-associated repeat expansion sites from Illumina PCR-free whole-genome sequencing data (Dolzhenko et al., 2019). ExpansionHunter estimates the number of copies of repeated short unit sequences by performing a targeted search through a BAM file for reads that span, flank, or are fully contained within each repeat. By combining evidence from multiple read signals, this approach is capable of genotyping repeats at a locus of interest even when the expanded repeat is substantially larger than the read length. Specifically, the method is capable of discovering and accurately estimating the size of *C9orf72* repeat expansions containing as many as ∼1000 (∼6Kbp) copies of the motif. For the *ATXN3*, heterozygous repeat calls were generated. The largest repeat allele for each sample was used for association with splicing and disease.

### Transcriptome-wide association study (TWAS)

We generated TWAS weights using FUSION (Gusev et al. 2016) for both gene expression and splicing, using the splice junction usage ratios computed by Leafcutter. The same spinal cord samples were used as in the QTL analysis, with the three spinal cord sections treated independently to create three splicing and three expression panels. For each reference panel, the same PEER covariates were used as for the QTL analysis. A linkage-disequilibrium matrix was created from the phased biallelic SNV and INDEL autosomal genetic variants (called *de novo* on the hg38 build (Lowy-Gallego et al. 2019)) for the selected subset of unrelated Non-Finnish European (NFE) samples (*n*=404) from the 1000 Genomes data (1000 Genomes Project Consortium et al. 2015) This custom LD reference data was then annotated with dbSNPv151 and the variants with HWE *P* < 10^−6^ were excluded. For the pre-computed weights we used an existing European LD reference mapped to the hg19 build (see **URLs**). TWAS uses GCTA-GREML (Yang et al. 2010) to estimate cis-SNP heritability (all SNPs 1Mbp from gene or intron start site) for each gene or splice junction. Only genes or splice junctions that were significant for heritability estimates at a Bonferroni-corrected p < 0.05 were retained for further analysis. Only variants present in the LD matrix were used to construct the weights. The gene expression or splice junction usage predictive weights were then computed by four different models implemented in the FUSION framework: best linear unbiased prediction, LASSO, Elastic Net, and top SNPs. The model with the best cross-validation prediction accuracy is then used to impute expression or splice junction usage into a GWAS, in this case the latest available ALS GWAS (Nicolas et al. 2018). The imputed gene expression or intron usage is then associated with disease risk, creating a Z-score and P-value. To account for multiple hypotheses, we applied an FDR of 5% within each expression and splicing reference panel. For comparison, pre-computed expression and splicing TWAS weights from 452 dorsolateral prefrontal cortex samples as part of the CommonMind Consortium (Li et al. 2019) were downloaded from the FUSION website (see URLs).

All plots were created using ggplot2 (Wickham 2009) in R (version 3.6.0), with ggrepel (Slowikowski, 2021), ggfortify (Tang, Horikoshi, and Li 2016), patchwork (Pedersen 2019), and ggbio (Yin, Cook, and Lawrence 2012) for additional layers of visualization.

## Supporting information

Supplementary Tables 1-14 and Supplementary Acknowledgments

## Data Availability

All raw RNA-seq data can be accessed via the NCBI's GEO database (GEO GSE137810, GSE124439, GSE116622, and GSE153960). Whole genome sequencing data will be uploaded to dbGaP upon publication. Full summary statistics for expression and splicing QTLs have been deposited on Zenodo (10.5281/zenodo.5248758). All TWAS weight files have been deposited on Zenodo (10.5281/zenodo.5256613). All RNA-seq and whole genome sequencing data generated by the NYGC ALS Consortium are made immediately available to all members of the Consortium and with other consortia with whom we have a reciprocal sharing arrangement. To request immediate access to new and ongoing data generated by the NYGC ALS Consortium and for samples provided through the Target ALS Postmortem Core, complete a genetic data request form at.

https://zenodo.org/record/5248758

https://github.com/jackhump/ALS_SpinalCord_QTLs

## Data availability

All raw RNA-seq data can be accessed via the NCBI’s GEO database (GEO GSE137810, GSE124439, GSE116622, and GSE153960). Whole genome sequencing data can be accessed via dbGAP (in progress). Full summary statistics for expression and splicing QTLs have been deposited on Zenodo (10.5281/zenodo.5248758). All TWAS weight files have been deposited on Zenodo (10.5281/zenodo.5256613). All RNA-seq and whole genome sequencing data generated by the NYGC ALS Consortium are made immediately available to all members of the Consortium and with other consortia with whom we have a reciprocal sharing arrangement. To request immediate access to new and ongoing data generated by the NYGC ALS Consortium and for samples provided through the Target ALS Postmortem Core, complete a genetic data request form at.

## Code availability

All analysis code written in R is available in Rmarkdown workbooks in a Github repository, and specific data processing pipelines are in separate repositories (**see URLs**).

## URLs

Code written for this project: https://github.com/jackhump/ALS_SpinalCord_QTLs

Full QTL summary statistics: https://zenodo.org/record/5248758

Full TWAS weights: https://doi.org/10.5281/zenodo.5256613

Molecular Signatures Database (MSigDb): http://www.gsea-msigdb.org/gsea/msigdb/index.jsp

Kelley et. al. gene fidelity marker genes: http://oldhamlab.ctec.ucsf.edu/data-download/

ENCODE Blacklist: https://github.com/Boyle-Lab/Blacklist/blob/master/lists/hg38-blacklist.v2.bed.gz

WGS QC pipeline: https://github.com/jackhump/WGS-QC-Pipeline

QTL mapping pipeline: https://github.com/RajLabMSSM/QTL-mapping-pipeline

DLPFC TWAS weights: http://gusevlab.org/projects/fusion/#reference-functional-data

ExpansionHunter: https://github.com/Illumina/ExpansionHunter

VCFs of 1000 Genomes samples: ftp://ftp.1000genomes.ebi.ac.uk/vol1/ftp/data_collections/1000_genomes_project/release/20190312_biallelic_SNV_and_INDEL/

## Author contributions

JH and TR conceived and designed the project. JH led the main analysis, with SV, RH, JTH, KPL, FK, KS, MBB, GN, USE contributing code and performing additional data analyses. JH and TR oversaw all aspects of the study, with input from DAK, HP and PF. DF and HP designed the sample collection methodology, reviewed sample and data quality, and coordinated NYGC ALS Consortium postmortem RNA research activity. The NYGC ALS Consortium and the Target ALS Human Postmortem Tissue Core provided human tissue samples as well as pathological, genetic, and clinical information. JH wrote the manuscript with input from all co-authors.

## Acknowledgements

We thank all members of the Raj lab for their feedback on the manuscript. JH, KPL and TR are funded by grants from the US National Institutes of Health (NIH NIA R56-AG055824 and NIA U01-AG068880). JTH is funded by an NIH Medical Scientist Training Program grant (T3GM007280). FK is supported by a BOF DOCPRO fellowship of the University of Antwerp Research Fund. PF is funded by the UK MRCl (MR/M008606/1 and MR/S006508/1), the UK Motor Neurone Disease Association, Rosetrees Trust and the UCLH NIHR Biomedical Research Centre. This work was supported in part through the computational resources and staff expertise provided by Scientific Computing at the Icahn School of Medicine at Mount Sinai. Research reported in this paper was supported by the Office of Research Infrastructure of the National Institutes of Health under award number S10OD018522 and S10OD026880. All NYGC ALS Consortium activities are supported by the ALS Association (ALSA, 19-SI-459) and the Tow Foundation. The funders had no role in study design, data collection and analysis, decision to publish or preparation of the manuscript.

## References

1000 Genomes Project Consortium, Adam Auton, Lisa D. Brooks, Richard M. Durbin, Erik P. Garrison, Hyun Min Kang, Jan O. Korbel, et al. 2015. “A Global Reference for Human Genetic Variation.” Nature 526 (7571): 68–74.

Adelson, Robert P., Alan E. Renton, Wentian Li, Nir Barzilai, Gil Atzmon, Alison M. Goate, Peter Davies, and Yun Freudenberg-Hua. 2019. “Empirical Design of a Variant Quality Control Pipeline for Whole Genome Sequencing Data Using Replicate Discordance.” Scientific Reports 9 (1): 16156.

Aguet, François, Alvaro N. Barbeira, Rodrigo Bonazzola, Andrew Brown, Stephane E. Castel, Brian Jo, Silva Kasela, et al. 2019. “The GTEx Consortium Atlas of Genetic Regulatory Effects across Human Tissues.” bioRxiv. https://doi.org/10.1101/787903.

Amemiya, Haley M., Anshul Kundaje, and Alan P. Boyle. 2019. “The ENCODE Blacklist: Identification of Problematic Regions of the Genome.” Scientific Reports 9 (1): 9354.

Andrés-Benito, Pol, Jesús Moreno, Ester Aso, Mónica Povedano, and Isidro Ferrer. 2017. “Amyotrophic Lateral Sclerosis, Gene Deregulation in the Anterior Horn of the Spinal Cord and Frontal Cortex Area 8: Implications in Frontotemporal Lobar Degeneration.” Aging 9 (3): 823–51.

Aspenström, Pontus. 2010. “Formin-Binding Proteins: Modulators of Formin-Dependent Actin Polymerization.” Biochimica et Biophysica Acta 1803 (2): 174–82.

Blum, Jacob A., Sandy Klemm, Jennifer L. Shadrach, Kevin A. Guttenplan, Lisa Nakayama, Arwa Kathiria, Phuong T. Hoang, et al. 2021. “Single-Cell Transcriptomic Analysis of the Adult Mouse Spinal Cord Reveals Molecular Diversity of Autonomic and Skeletal Motor Neurons.” Nature Neuroscience 24 (4): 572–83.

Boillée, Séverine, Koji Yamanaka, Christian S. Lobsiger, Neal G. Copeland, Nancy A. Jenkins, George Kassiotis, George Kollias, and Don W. Cleveland. 2006. “Onset and Progression in Inherited ALS Determined by Motor Neurons and Microglia.” Science 312 (5778): 1389–92.

Bolduc, Benjamin. n.d. “Quality Control of Reads Using Trimmomatic (Cyverse) v1 (protocols.io.ewbbfan).” Protocols.io. https://doi.org/10.17504/protocols.io.ewbbfan.

Brettschneider, Johannes, Jon B. Toledo, Vivianna M. Van Deerlin, Lauren Elman, Leo McCluskey, Virginia M-Y Lee, and John Q. Trojanowski. 2012. “Microglial Activation Correlates with Disease Progression and Upper Motor Neuron Clinical Symptoms in Amyotrophic Lateral Sclerosis.” PloS One 7 (6): e39216.

Brohawn, David G., Laura C. O’Brien, and James P. Bennett Jr. 2016. “RNAseq Analyses Identify Tumor Necrosis Factor-Mediated Inflammation as a Major Abnormality in ALS Spinal Cord.” PloS One 11 (8): e0160520.

Byrne, Susan, Cathal Walsh, Catherine Lynch, Peter Bede, Marwa Elamin, Kevin Kenna, Russell McLaughlin, and Orla Hardiman. 2011. “Rate of Familial Amyotrophic Lateral Sclerosis: A Systematic Review and Meta-Analysis.” Journal of Neurology, Neurosurgery, and Psychiatry 82 (6): 623–27.

Chang, Christopher C., Carson C. Chow, Laurent Cam Tellier, Shashaank Vattikuti, Shaun M. Purcell, and James J. Lee. 2015. “Second-Generation PLINK: Rising to the Challenge of Larger and Richer Datasets.” GigaScience. https://doi.org/10.1186/s13742-015-0047-8.

Chen, Wei-Ting, Ashley Lu, Katleen Craessaerts, Benjamin Pavie, Carlo Sala Frigerio, Nikky Corthout, Xiaoyan Qian, et al. 2020. “Spatial Transcriptomics and In Situ Sequencing to Study Alzheimer’s Disease.” Cell 182 (4): 976–91.e19.

Chiu, Isaac M., Emiko T. A. Morimoto, Hani Goodarzi, Jennifer T. Liao, Sean O’Keeffe, Hemali P. Phatnani, Michael Muratet, et al. 2013. “A Neurodegeneration-Specific Gene-Expression Signature of Acutely Isolated Microglia from an Amyotrophic Lateral Sclerosis Mouse Model.” Cell Reports 4 (2): 385–401.

Cirulli, Elizabeth T., Brittany N. Lasseigne, Slavé Petrovski, P. C. Sapp, Patrick A. Dion, C. S. Leblond, Julien Couthouis, et al. 2016. “Exome Sequencing in Amyotrophic Lateral Sclerosis Identifies Risk Genes and Pathways.” Nature Methods 347 (6229): 1436–41.

Consortium, The Gtex, and The GTEx Consortium. 2020. “The GTEx Consortium Atlas of Genetic Regulatory Effects across Human Tissues.” Science. https://doi.org/10.1126/science.aaz1776.

Danecek, Petr, Adam Auton, Goncalo Abecasis, Cornelis A. Albers, Eric Banks, Mark A. DePristo, Robert E. Handsaker, et al. 2011. “The Variant Call Format and VCFtools.” Bioinformatics 27 (15): 2156–58.

Danecek, Petr, James K. Bonfield, Jennifer Liddle, John Marshall, Valeriu Ohan, Martin O. Pollard, Andrew Whitwham, et al. 2021. “Twelve Years of SAMtools and BCFtools.” GigaScience 10 (2). https://doi.org/10.1093/gigascience/giab008.

Darmanis, Spyros, Steven A. Sloan, Ye Zhang, Martin Enge, Christine Caneda, Lawrence M. Shuer, Melanie G. Hayden Gephart, Ben A. Barres, and Stephen R. Quake. 2015. “A Survey of Human Brain Transcriptome Diversity at the Single Cell Level.” Proceedings of the National Academy of Sciences. https://doi.org/10.1073/pnas.1507125112.

DeJesus-Hernandez, Mariely, Ian R. Mackenzie, Bradley F. Boeve, Adam L. Boxer, Matt Baker, Nicola J. Rutherford, Alexandra M. Nicholson, et al. 2011. “Expanded GGGGCC Hexanucleotide Repeat in Noncoding Region of C9ORF72 Causes Chromosome 9p-Linked FTD and ALS.” Neuron 72 (2): 245–56.

D’Erchia, Anna Maria, Angela Gallo, Caterina Manzari, Susanna Raho, David S. Horner, Matteo Chiara, Alessio Valletti, et al. 2017. “Massive Transcriptome Sequencing of Human Spinal Cord Tissues Provides New Insights into Motor Neuron Degeneration in ALS.” Scientific Reports. https://doi.org/10.1038/s41598-017-10488-7.

Dickson, Dennis W., Matthew C. Baker, Jazmyne L. Jackson, Mariely DeJesus-Hernandez, Nicole A. Finch, Shulan Tian, Michael G. Heckman, et al. 2019. “Extensive Transcriptomic Study Emphasizes Importance of Vesicular Transport in C9orf72 Expansion Carriers.” Acta Neuropathologica Communications 7 (1): 150.

Di Tommaso, Paolo, Maria Chatzou, Evan W. Floden, Pablo Prieto Barja, Emilio Palumbo, and Cedric Notredame. 2017. “Nextflow Enables Reproducible Computational Workflows.” Nature Biotechnology 35 (4): 316–19.

Dobin, Alexander, Carrie A. Davis, Felix Schlesinger, Jorg Drenkow, Chris Zaleski, Sonali Jha, Philippe Batut, Mark Chaisson, and Thomas R. Gingeras. 2013. “STAR: Ultrafast Universal RNA-Seq Aligner.” Bioinformatics 29 (1): 15–21.

Dols-Icardo, Oriol, VÍctor Montal, Sònia Sirisi, Gema López-Pernas, Laura Cervera-Carles, Marta Querol-Vilaseca, Laia Muñoz, et al. 2020. “Motor Cortex Transcriptome Reveals Microglial Key Events in Amyotrophic Lateral Sclerosis.” Neurology(R) Neuroimmunology & Neuroinflammation 7 (5). https://doi.org/10.1212/NXI.0000000000000829.

Dolzhenko, Egor, Viraj Deshpande, Felix Schlesinger, Peter Krusche, Roman Petrovski, Sai Chen, Dorothea Emig-Agius, et al. 2019. “ExpansionHunter: A Sequence-Graph Based Tool to Analyze Variation in Short Tandem Repeat Regions.” https://doi.org/10.1101/572545.

Efthymiou, Stephanie, Vincenzo Salpietro, Nancy Malintan, Mallory Poncelet, Yamna Kriouile, Sara Fortuna, Rita De Zorzi, et al. 2019. “Biallelic Mutations in Neurofascin Cause Neurodevelopmental Impairment and Peripheral Demyelination.” Brain: A Journal of Neurology 142 (10): 2948–64.

Elden, Andrew C., Hyung Jun Kim, Michael P. Hart, Alice S. Chen-Plotkin, Brian S. Johnson, Xiaodong Fang, Maria Armakola, et al. 2010. “Ataxin-2 Intermediate-Length Polyglutamine Expansions Are Associated with Increased Risk for ALS.” Nature 466 (7310): 1069–75.

Es, Michael A. van, Jan H. Veldink, Christiaan G. J. Saris, Hylke M. Blauw, Paul W. J. van Vught, Anna Birve, Robin Lemmens, et al. 2009. “Genome-Wide Association Study Identifies 19p13.3 (UNC13A) and 9p21.2 as Susceptibility Loci for Sporadic Amyotrophic Lateral Sclerosis.” Nature Genetics 41 (10): 1083–87.

Ewels, Philip, Måns Magnusson, Sverker Lundin, and Max Käller. 2016. “MultiQC: Summarize Analysis Results for Multiple Tools and Samples in a Single Report.” Bioinformatics 32 (19): 3047–48.

Feng, Yang-Yang, Avinash Ramu, Kelsy C. Cotto, Zachary L. Skidmore, Jason Kunisaki, Donald F. Conrad, Yiing Lin, et al. 2018. “RegTools: Integrated Analysis of Genomic and Transcriptomic Data for Discovery of Splicing Variants in Cancer.” bioRxiv. https://doi.org/10.1101/436634.

Fort, Alexandre, Nikolaos I. Panousis, Marco Garieri, Stylianos E. Antonarakis, Tuuli Lappalainen, Emmanouil T. Dermitzakis, and Olivier Delaneau. 2017. “MBV: A Method to Solve Sample Mislabeling and Detect Technical Bias in Large Combined Genotype and Sequencing Assay Datasets.” Bioinformatics 33 (12): 1895–97.

Giambartolomei, Claudia, Damjan Vukcevic, Eric E. Schadt, Lude Franke, Aroon D. Hingorani, Chris Wallace, and Vincent Plagnol. 2014. “Bayesian Test for Colocalisation between Pairs of Genetic Association Studies Using Summary Statistics.” PLoS Genetics 10 (5): e1004383.

Gusev, Alexander, Arthur Ko, Huwenbo Shi, Gaurav Bhatia, Wonil Chung, Brenda W. J. H. Penninx, Rick Jansen, et al. 2016. “Integrative Approaches for Large-Scale Transcriptome-Wide Association Studies.” Nature Genetics 48 (3): 245–52.

Guttenplan, Kevin A., Maya K. Weigel, Drew I. Adler, Julien Couthouis, Shane A. Liddelow, Aaron D. Gitler, and Ben A. Barres. 2020. “Knockout of Reactive Astrocyte Activating Factors Slows Disease Progression in an ALS Mouse Model.” Nature Communications 11 (1): 3753.

Habib, Naomi, Cristin McCabe, Sedi Medina, Miriam Varshavsky, Daniel Kitsberg, Raz Dvir-Szternfeld, Gilad Green, et al. 2020. “Disease-Associated Astrocytes in Alzheimer’s Disease and Aging.” Nature Neuroscience 23 (6): 701–6.

Haidet-Phillips, Amanda M., Mark E. Hester, Carlos J. Miranda, Kathrin Meyer, Lyndsey Braun, Ashley Frakes, Sungwon Song, et al. 2011. “Astrocytes from Familial and Sporadic ALS Patients Are Toxic to Motor Neurons.” Nature Biotechnology 29 (9): 824–28.

Harrow, Jennifer, Adam Frankish, Jose M. Gonzalez, and Kelly A. Frazer. 2012. “GENCODE : The Reference Human Genome Annotation for The ENCODE Project.” Genome Research 22: 1760–74.

Hirano, Makito, Makoto Samukawa, Chiharu Isono, Kazumasa Saigoh, Yusaku Nakamura, and Susumu Kusunoki. 2018. “Noncoding Repeat Expansions for ALS in Japan Are Associated with the ATXN8OS Gene.” Neurology Genetics. https://doi.org/10.1212/nxg.0000000000000252.

Hoffman, Gabriel E., and Eric E. Schadt. 2016. “variancePartition: Interpreting Drivers of Variation in Complex Gene Expression Studies.” BMC Bioinformatics 17 (1): 483.

Ho, Ritchie, Michael J. Workman, Pranav Mathkar, Kathryn Wu, Kevin J. Kim, Jacqueline G. O’Rourke, Mariko Kellogg, et al. 2021. “Cross-Comparison of Human iPSC Motor Neuron Models of Familial and Sporadic ALS Reveals Early and Convergent Transcriptomic Disease Signatures.” Cell Systems 12 (2): 159–75.e9.

Hunt, Gregory J., Saskia Freytag, Melanie Bahlo, and Johann A. Gagnon-Bartsch. 2019. “Dtangle: Accurate and Robust Cell Type Deconvolution.” Bioinformatics 35 (12): 2093–99.

Hüttenrauch, Melanie, Isabella Ogorek, Hans Klafki, Markus Otto, Christine Stadelmann, Sascha Weggen, Jens Wiltfang, and Oliver Wirths. 2018. “Glycoprotein NMB: A Novel Alzheimer’s Disease Associated Marker Expressed in a Subset of Activated Microglia.” Acta Neuropathologica Communications 6 (1): 108.

Jaarsma, Dick, Eva Teuling, Elize D. Haasdijk, Chris I. De Zeeuw, and Casper C. Hoogenraad. 2008. “Neuron-Specific Expression of Mutant Superoxide Dismutase Is Sufficient to Induce Amyotrophic Lateral Sclerosis in Transgenic Mice.” The Journal of Neuroscience: The Official Journal of the Society for Neuroscience 28 (9): 2075–88.

Jackson, Jazmyne L., Nicole A. Finch, Matthew C. Baker, Jennifer M. Kachergus, Mariely DeJesus-Hernandez, Kimberly Pereira, Elizabeth Christopher, et al. 2020. “Elevated Methylation Levels, Reduced Expression Levels, and Frequent Contractions in a Clinical Cohort of C9orf72 Expansion Carriers.” Molecular Neurodegeneration. https://doi.org/10.1186/s13024-020-0359-8.

Kang, Shin H., Ying Li, Masahiro Fukaya, Ileana Lorenzini, Don W. Cleveland, Lyle W. Ostrow, Jeffrey D. Rothstein, and Dwight E. Bergles. 2013. “Degeneration and Impaired Regeneration of Gray Matter Oligodendrocytes in Amyotrophic Lateral Sclerosis.” Nature Neuroscience 16 (5): 571–79.

Kelley, Kevin W., Hiromi Nakao-Inoue, Anna V. Molofsky, and Michael C. Oldham. 2018. “Variation among Intact Tissue Samples Reveals the Core Transcriptional Features of Human CNS Cell Classes.” Nature Neuroscience 21 (9): 1171–84.

Kenna, Kevin P., Perry T. C. van Doormaal, Annelot M. Dekker, Nicola Ticozzi, Brendan J. Kenna, Frank P. Diekstra, Wouter van Rheenen, et al. 2016. “NEK1 Variants Confer Susceptibility to Amyotrophic Lateral Sclerosis.” Nature Genetics. https://doi.org/10.1038/ng.3626.

Keren-Shaul, Hadas, Amit Spinrad, Assaf Weiner, Orit Matcovitch-Natan, Raz Dvir-Szternfeld, Tyler K. Ulland, Eyal David, et al. 2017. “A Unique Microglia Type Associated with Restricting Development of Alzheimer’s Disease.” Cell 169 (7): 1276–90.e17.

Kim-Hellmuth, Sarah, François Aguet, Meritxell Oliva, Manuel Muñoz-Aguirre, Silva Kasela, Valentin Wucher, Stephane E. Castel, et al. 2020. “Cell Type-Specific Genetic Regulation of Gene Expression across Human Tissues.” Science 369 (6509). https://doi.org/10.1126/science.aaz8528.

Klemens, J., M. Ciurkiewicz, E. Chludzinski, M. Iseringhausen, D. Klotz, V. M. Pfankuche, R. Ulrich, et al. 2019. “Neurotoxic Potential of Reactive Astrocytes in Canine Distemper Demyelinating Leukoencephalitis.” Scientific Reports 9 (1): 11689.

Klim, Joseph R., Luis A. Williams, Francesco Limone, Irune Guerra San Juan, Brandi N. Davis-Dusenbery, Daniel A. Mordes, Aaron Burberry, et al. 2019. “ALS-Implicated Protein TDP-43 Sustains Levels of STMN2, a Mediator of Motor Neuron Growth and Repair.” Nature Neuroscience 22 (2): 167–79.

Köster, Johannes, and Sven Rahmann. 2012. “Snakemake-a Scalable Bioinformatics Workflow Engine.” Bioinformatics 28 (19): 2520–22.

Lake, Blue B., Rizi Ai, Gwendolyn E. Kaeser, Neeraj S. Salathia, Yun C. Yung, Rui Liu, Andre Wildberg, et al. 2016. “Neuronal Subtypes and Diversity Revealed by Single-Nucleus RNA Sequencing of the Human Brain.” Science 352 (6293): 1586–90.

Lattante, Serena, Maria Grazia Pomponi, Amelia Conte, Giuseppe Marangi, Giulia Bisogni, Agata Katia Patanella, Emiliana Meleo, et al. 2018. “ATXN1 Intermediate-Length Polyglutamine Expansions Are Associated with Amyotrophic Lateral Sclerosis.” Neurobiology of Aging 64 (April): 157.e1–157.e5.

Law, Charity W., Yunshun Chen, Wei Shi, and Gordon K. Smyth. 2014. “Voom: Precision Weights Unlock Linear Model Analysis Tools for RNA-Seq Read Counts.” Genome Biology 15 (2): R29.

Lepore, Angelo C., Britta Rauck, Christine Dejea, Andrea C. Pardo, Mahendra S. Rao, Jeffrey D. Rothstein, and Nicholas J. Maragakis. 2008. “Focal Transplantation–based Astrocyte Replacement Is Neuroprotective in a Model of Motor Neuron Disease.” Nature Neuroscience 11 (11): 1294–1301.

Liberzon, Arthur, Chet Birger, Helga Thorvaldsdóttir, Mahmoud Ghandi, Jill P. Mesirov, and Pablo Tamayo. 2015. “The Molecular Signatures Database (MSigDB) Hallmark Gene Set Collection.” Cell Systems 1 (6): 417–25.

Li, Bo, and Colin N. Dewey. 2011. “RSEM: Accurate Transcript Quantification from RNA-Seq Data with or without a Reference Genome.” BMC Bioinformatics 12 (August): 323.

Liddelow, Shane A., Kevin A. Guttenplan, Laura E. Clarke, Frederick C. Bennett, Christopher J. Bohlen, Lucas Schirmer, Mariko L. Bennett, et al. 2017. “Neurotoxic Reactive Astrocytes Are Induced by Activated Microglia.” Nature 541 (7638): 481–87.

Li, Heng, Bob Handsaker, Alec Wysoker, Tim Fennell, Jue Ruan, Nils Homer, Gabor Marth, Goncalo Abecasis, Richard Durbin, and 1000 Genome Project Data Processing Subgroup. 2009. “The Sequence Alignment/Map Format and SAMtools.” Bioinformatics 25 (16): 2078–79.

Li, Yang I., David A. Knowles, Jack Humphrey, Alvaro N. Barbeira, Scott P. Dickinson, Hae Kyung Im, and Jonathan K. Pritchard. 2018. “Annotation-Free Quantification of RNA Splicing Using LeafCutter.” Nature Genetics 50 (1): 151–58.

Li, Yang I., Garrett Wong, Jack Humphrey, and Towfique Raj. 2019. “Prioritizing Parkinson’s Disease Genes Using Population-Scale Transcriptomic Data.” Nature Communications 10 (1): 994.

Lopes, Katia de Paiva, Katia de Paiva Lopes, Gijsje J. L. Snijders, Jack Humphrey, Amanda Allan, Marjolein Sneeboer, Elisa Navarro, et al. n.d. “Atlas of Genetic Effects in Human Microglia Transcriptome across Brain Regions, Aging and Disease Pathologies.” https://doi.org/10.1101/2020.10.27.356113.

Love, Michael I., Charlotte Soneson, and Mark D. Robinson. 2017. “Importing Transcript Abundance Datasets with Tximport.” Dim (txi. Inf. Rep $ infReps $ sample1) 1 (178136): 5.

Lowy-Gallego, Ernesto, Susan Fairley, Xiangqun Zheng-Bradley, Magali Ruffier, Laura Clarke, Paul Flicek, and 1000 Genomes Project Consortium. 2019. “Variant Calling on the GRCh38 Assembly with the Data from Phase Three of the 1000 Genomes Project.” Wellcome Open Research 4 (December): 50.

Majounie, Elisa, Alan E. Renton, Kin Mok, Elise G. P. Dopper, Adrian Waite, Sara Rollinson, Adriano Chiò, et al. 2012. “Frequency of the C9orf72 Hexanucleotide Repeat Expansion in Patients with Amyotrophic Lateral Sclerosis and Frontotemporal Dementia: A Cross-Sectional Study.” Lancet Neurology 11 (4): 323–30.

Månberg, Anna, Nathan Skene, Folkert Sanders, Marta Trusohamn, Julia Remnestål, Anna Szczepińska, Inci Sevval Aksoylu, et al. 2021. “Publisher Correction: Altered Perivascular Fibroblast Activity Precedes ALS Disease Onset.” Nature Medicine 27 (7): 1308.

Mancarci, O., and L. French. 2019. “Homologene: Quick Access to Homologene and Gene Annotation Updates.” R Package Version 1: 68.

Maniatis, Silas, Tarmo Äijö, Sanja Vickovic, Catherine Braine, Kristy Kang, Annelie Mollbrink, Delphine Fagegaltier, et al. 2019. “Spatiotemporal Dynamics of Molecular Pathology in Amyotrophic Lateral Sclerosis.” Science 364 (6435): 89–93.

Manichaikul, Ani, Josyf C. Mychaleckyj, Stephen S. Rich, Kathy Daly, Michèle Sale, and Wei-Min Chen. 2010. “Robust Relationship Inference in Genome-Wide Association Studies.” Bioinformatics 26 (22): 2867–73.

Mathys, Hansruedi, Jose Davila-Velderrain, Zhuyu Peng, Fan Gao, Shahin Mohammadi, Jennie Z. Young, Madhvi Menon, et al. 2019. “Single-Cell Transcriptomic Analysis of Alzheimer’s Disease.” Nature 570 (7761): 332–37.

Melamed, Ze’ev, Jone López-Erauskin, Michael W. Baughn, Ouyang Zhang, Kevin Drenner, Ying Sun, Fernande Freyermuth, et al. 2019. “Premature Polyadenylation-Mediated Loss of Stathmin-2 Is a Hallmark of TDP-43-Dependent Neurodegeneration.” Nature Neuroscience 22 (2): 180–90.

Murthy, Megha N., Cornelis Blauwendraat, UKBEC, Sebastian Guelfi, IPDGC, John Hardy, Patrick A. Lewis, and Daniah Trabzuni. 2017. “Increased Brain Expression of GPNMB Is Associated with Genome Wide Significant Risk for Parkinson’s Disease on Chromosome 7p15.3.” Neurogenetics 18 (3): 121–33.

Myers, Timothy A., Stephen J. Chanock, and Mitchell J. Machiela. 2020. “LDlinkR: An R Package for Rapidly Calculating Linkage Disequilibrium Statistics in Diverse Populations.” Frontiers in Genetics 11 (February): 157.

Nakamura, Ryoichi, Kazuharu Misawa, Genki Tohnai, Masahiro Nakatochi, Sho Furuhashi, Naoki Atsuta, Naoki Hayashi, et al. 2020. “A Multi-Ethnic Meta-Analysis Identifies Novel Genes, Including ACSL5, Associated with Amyotrophic Lateral Sclerosis.” Communications Biology 3 (1): 526.

Nalls, Mike A., Cornelis Blauwendraat, Costanza L. Vallerga, Karl Heilbron, Sara Bandres-Ciga, Diana Chang, Manuela Tan, et al. 2019. “Identification of Novel Risk Loci, Causal Insights, and Heritable Risk for Parkinson’s Disease: A Meta-Analysis of Genome-Wide Association Studies.” Lancet Neurology 18 (12): 1091–1102.

Nelson, Andrew D., and Paul M. Jenkins. 2017. “Axonal Membranes and Their Domains: Assembly and Function of the Axon Initial Segment and Node of Ranvier.” Frontiers in Cellular Neuroscience 11 (May): 136.

Neumann, M., D. M. Sampathu, L. K. Kwong, A. C. Truax, M. C. Micsenyi, T. T. Chou, J. Bruce, et al. 2006. “Ubiquitinated TDP-43 in Frontotemporal Lobar Degeneration and Amyotrophic Lateral Sclerosis.” Science 314 (5796): 130–33.

Nicolas, Aude, Kevin P. Kenna, Alan E. Renton, Nicola Ticozzi, Faraz Faghri, Ruth Chia, Janice A. Dominov, et al. 2018. “Genome-Wide Analyses Identify KIF5A as a Novel ALS Gene.” Neuron 97 (6): 1268–83.e6.

Novikova, Gloriia, Manav Kapoor, Julia Tcw, Edsel M. Abud, Anastasia G. Efthymiou, Steven X. Chen, Haoxiang Cheng, et al. 2021. “Integration of Alzheimer’s Disease Genetics and Myeloid Genomics Identifies Disease Risk Regulatory Elements and Genes.” Nature Communications 12 (1): 1610.

Oldham, Michael C., Genevieve Konopka, Kazuya Iwamoto, Peter Langfelder, Tadafumi Kato, Steve Horvath, and Daniel H. Geschwind. 2008. “Functional Organization of the Transcriptome in Human Brain.” Nature Neuroscience 11 (11): 1271–82.

O’Rourke, J. G., L. Bogdanik, A. Yáñez, D. Lall, A. J. Wolf, A. K. M. G. Muhammad, R. Ho, et al. 2016. “C9orf72 Is Required for Proper Macrophage and Microglial Function in Mice.” Science 351 (6279): 1324–29.

Oscanoa, Jorge, Lavanya Sivapalan, Emanuela Gadaleta, Abu Z. Dayem Ullah, Nicholas R. Lemoine, and Claude Chelala. 2020. “SNPnexus: A Web Server for Functional Annotation of Human Genome Sequence Variation (2020 Update).” Nucleic Acids Research 48 (W1): W185–92.

Paulson, Henry. 2006. “Machado-Joseph Disease/Spinocerebellar Ataxia Type 3.” Genetic Instabilities and Neurological Diseases. https://doi.org/10.1016/b978-012369462-1/50025-9.

Pedersen, T. L. 2019. “Patchwork: The Composer of Plots.” R Package Version 1 (0): 410.

Phatnani, Hemali P., Paolo Guarnieri, Brad A. Friedman, Monica A. Carrasco, Michael Muratet, Sean O’Keeffe, Chiamaka Nwakeze, et al. 2013. “Intricate Interplay between Astrocytes and Motor Neurons in ALS.” Proceedings of the National Academy of Sciences of the United States of America 110 (8): E756–65.

Poplin, Ryan, Valentin Ruano-Rubio, Mark A. DePristo, Tim J. Fennell, Mauricio O. Carneiro, Geraldine A. Van der Auwera, David E. Kling, et al. n.d. “Scaling Accurate Genetic Variant Discovery to Tens of Thousands of Samples.” https://doi.org/10.1101/201178.

Pramatarova, A., J. Laganière, J. Roussel, K. Brisebois, and G. A. Rouleau. 2001. “Neuron-Specific Expression of Mutant Superoxide Dismutase 1 in Transgenic Mice Does Not Lead to Motor Impairment.” The Journal of Neuroscience: The Official Journal of the Society for Neuroscience 21 (10): 3369–74.

Price, Alkes L., Nick J. Patterson, Robert M. Plenge, Michael E. Weinblatt, Nancy A. Shadick, and David Reich. 2006. “Principal Components Analysis Corrects for Stratification in Genome-Wide Association Studies.” Nature Genetics 38 (8): 904–9.

Prudencio, Mercedes, Veronique V. Belzil, Ranjan Batra, Christian A. Ross, Tania F. Gendron, Luc J. Pregent, Melissa E. Murray, et al. 2015. “Distinct Brain Transcriptome Profiles in C9orf72-Associated and Sporadic ALS.” Nature Neuroscience 18 (8): 1175–82.

Prudencio, Mercedes, Hector Garcia-Moreno, Karen R. Jansen-West, Rana Hanna Al-Shaikh, Tania F. Gendron, Michael G. Heckman, Matthew R. Spiegel, et al. 2020. “Toward Allele-Specific Targeting Therapy and Pharmacodynamic Marker for Spinocerebellar Ataxia Type 3.” Science Translational Medicine 12 (566). https://doi.org/10.1126/scitranslmed.abb7086.

Prudencio, Mercedes, Jack Humphrey, Sarah Pickles, Anna-Leigh Brown, Sarah E. Hill, Jennifer Kachergus, Ji Shi, et al. 2020. “Truncated Stathmin-2 Is a Marker of TDP-43 Pathology in Frontotemporal Dementia.” The Journal of Clinical Investigation, August. https://doi.org/10.1172/JCI139741.

Ravits, John M., and Albert R. La Spada. 2009. “ALS Motor Phenotype Heterogeneity, Focality, and Spread: Deconstructing Motor Neuron Degeneration.” Neurology 73 (10): 805–11.

Regier, Allison A., Yossi Farjoun, David E. Larson, Olga Krasheninina, Hyun Min Kang, Daniel P. Howrigan, Bo-Juen Chen, et al. 2018. “Functional Equivalence of Genome Sequencing Analysis Pipelines Enables Harmonized Variant Calling across Human Genetics Projects.” Nature Communications 9 (1): 4038.

Reimand, Jüri, Meelis Kull, Hedi Peterson, Jaanus Hansen, and Jaak Vilo. 2007. “g:Profiler—a Web-Based Toolset for Functional Profiling of Gene Lists from Large-Scale Experiments.” Nucleic Acids Research 35 (suppl_2): W193–200.

Renton, Alan E., Adriano Chiò, and Bryan J. Traynor. 2014. “State of Play in Amyotrophic Lateral Sclerosis Genetics.” Nature Neuroscience 17 (1): 17–23.

Renton, Alan E., Elisa Majounie, Adrian Waite, Javier Simón-Sánchez, Sara Rollinson, J. Raphael Gibbs, Jennifer C. Schymick, et al. 2011. “A Hexanucleotide Repeat Expansion in C9ORF72 Is the Cause of Chromosome 9p21-Linked ALS-FTD.” Neuron 72 (2): 257–68.

Rheenen, Wouter van, Rick A. A. van der Spek, Mark K. Bakker, Joke J. F. A. van Vugt, Paul J. Hop, Ramona A. J. Zwamborn, Niek de Klein, et al. 2021. “Common and Rare Variant Association Analyses in Amyotrophic Lateral Sclerosis Identify 15 Risk Loci with Distinct Genetic Architectures and Neuron-Specific Biology.” bioRxiv. medRxiv. https://doi.org/10.1101/2021.03.12.21253159.

Risso, Davide, Katja Schwartz, Gavin Sherlock, and Sandrine Dudoit. 2011. “GC-Content Normalization for RNA-Seq Data.” BMC Bioinformatics 12 (December): 480.

Saez-Atienzar, Sara, Sara Bandres-Ciga, Rebekah G. Langston, Jonggeol J. Kim, Shing Wan Choi, Regina H. Reynolds, the International ALS Genomics Consortium; ITALSGEN, et al. 2020. “Genetic Analysis of Amyotrophic Lateral Sclerosis Identifies Contributing Pathways and Cell Types.” Cold Spring Harbor Laboratory. https://doi.org/10.1101/2020.07.20.211276.

Saul, J., E. Hutchins, R. Reiman, M. Saul, L. W. Ostrow, B. T. Harris, K. Van Keuren-Jensen, R. Bowser, and N. Bakkar. 2020. “Global Alterations to the Choroid Plexus Blood-CSF Barrier in Amyotrophic Lateral Sclerosis.” Acta Neuropathologica Communications 8 (1): 92.

Schroeder, Andreas, Odilo Mueller, Susanne Stocker, Ruediger Salowsky, Michael Leiber, Marcus Gassmann, Samar Lightfoot, Wolfram Menzel, Martin Granzow, and Thomas Ragg. 2006. “The RIN: An RNA Integrity Number for Assigning Integrity Values to RNA Measurements.” BMC Molecular Biology 7 (January): 3.

Seidel, Kay, Wilfred F. A. den Dunnen, Christian Schultz, Henry Paulson, Stefanie Frank, Rob A. de Vos, Ewout R. Brunt, Thomas Deller, Harm H. Kampinga, and Udo Rüb. 2010. “Axonal Inclusions in Spinocerebellar Ataxia Type 3.” Acta Neuropathologica 120 (4): 449–60.

Skene, Nathan G., and Seth G. N. Grant. 2016. “Identification of Vulnerable Cell Types in Major Brain Disorders Using Single Cell Transcriptomes and Expression Weighted Cell Type Enrichment.” Frontiers in Neuroscience 10 (January): 16.

Slowikowski, Kamil. n.d. “Ggrepel: Repulsive Text and Label Geoms for ‘ggplot2’, 2016.” R Package Version 0. 5.

Stegle, Oliver, Leopold Parts, Matias Piipari, John Winn, and Richard Durbin. 2012. “Using Probabilistic Estimation of Expression Residuals (PEER) to Obtain Increased Power and Interpretability of Gene Expression Analyses.” Nature Protocols 7 (3): 500.

Storey, John D. 2003. “The Positive False Discovery Rate: A Bayesian Interpretation and the Q-Value.” Annals of Statistics 31 (6): 2013–35.

Subramanian, Aravind, Pablo Tamayo, Vamsi K. Mootha, Sayan Mukherjee, Benjamin L. Ebert, Michael A. Gillette, Amanda Paulovich, et al. 2005. “Gene Set Enrichment Analysis: A Knowledge-Based Approach for Interpreting Genome-Wide Expression Profiles.” Proceedings of the National Academy of Sciences of the United States of America 102 (43): 15545–50.

Tam, Oliver H., Nikolay V. Rozhkov, Regina Shaw, Duyang Kim, Isabel Hubbard, Samantha Fennessey, Nadia Propp, et al. 2019. “Postmortem Cortex Samples Identify Distinct Molecular Subtypes of ALS: Retrotransposon Activation, Oxidative Stress, and Activated Glia.” Cell Reports 29 (5): 1164–77.e5.

Tanaka, Hirotaka, Masamitsu Shimazawa, Masataka Kimura, Masafumi Takata, Kazuhiro Tsuruma, Mitsunori Yamada, Hitoshi Takahashi, et al. 2012. “The Potential of GPNMB as Novel Neuroprotective Factor in Amyotrophic Lateral Sclerosis.” Scientific Reports 2 (August): 573.

Tang, Yuan, Masaaki Horikoshi, and Wenxuan Li. 2016. “Ggfortify: Unified Interface to Visualize Statistical Results of Popular R Packages.” The R Journal 8 (2): 474.

Taylor-Weiner, Amaro, François Aguet, Nicholas J. Haradhvala, Sager Gosai, Shankara Anand, Jaegil Kim, Kristin Ardlie, Eliezer M. Van Allen, and Gad Getz. 2019. “Scaling Computational Genomics to Millions of Individuals with GPUs.” Genome Biology 20 (1): 228.

Tazelaar, Gijs H. P., Steven Boeynaems, Mathias De Decker, Joke J. F. A. van Vugt, Lindy Kool, H. Stephan Goedee, Russell L. McLaughlin, et al. 2020. “Repeat Expansions Confer Risk for Amyotrophic Lateral Sclerosis and Contribute to TDP-43 Mislocalization.” Brain Communications 2 (2): fcaa064.

Thompson, Alexander G., Elizabeth Gray, Marie-Laëtitia Thézénas, Philip D. Charles, Samuel Evetts, Michele T. Hu, Kevin Talbot, Roman Fischer, Benedikt M. Kessler, and Martin R. Turner. 2018. “Cerebrospinal Fluid Macrophage Biomarkers in Amyotrophic Lateral Sclerosis.” Annals of Neurology 83 (2): 258–68.

Town, Terrence, Veljko Nikolic, and Jun Tan. 2005. “The Microglial ‘Activation’ Continuum: From Innate to Adaptive Responses.” Journal of Neuroinflammation 2 (1): 24.

Van Rheenen, Wouter, Aleksey Shatunov, Annelot M. Dekker, Russell L. McLaughlin, Frank P. Diekstra, Sara L. Pulit, Rick A. A. Van Der Spek, et al. 2016. “Genome-Wide Association Analyses Identify New Risk Variants and the Genetic Architecture of Amyotrophic Lateral Sclerosis.” Nature Genetics 48 (9): 1043–48.

Wainberg, Michael, Nasa Sinnott-Armstrong, Nicholas Mancuso, Alvaro N. Barbeira, David A. Knowles, David Golan, Raili Ermel, et al. 2019. “Opportunities and Challenges for Transcriptome-Wide Association Studies.” Nature Genetics 51 (4): 592–99.

Wang, Lijun, Kamal Sharma, Gabriella Grisotti, and Raymond P. Roos. 2009. “The Effect of Mutant SOD1 Dismutase Activity on Non-Cell Autonomous Degeneration in Familial Amyotrophic Lateral Sclerosis.” Neurobiology of Disease 35 (2): 234–40.

Wang, Xuran, Jihwan Park, Katalin Susztak, Nancy R. Zhang, and Mingyao Li. 2019. “Bulk Tissue Cell Type Deconvolution with Multi-Subject Single-Cell Expression Reference.” Nature Communications 10 (1): 380.

West, Ryan J. H., Chris Ugbode, Fen-Biao Gao, and Sean T. Sweeney. 2018. “The pro-Apoptotic JNK Scaffold POSH/SH3RF1 Mediates CHMP2BIntron5-Associated Toxicity in Animal Models of Frontotemporal Dementia.” Human Molecular Genetics 27 (8): 1382–95.

Wickham, Hadley. 2009. ggplot2: Elegant Graphics for Data Analysis. Springer-Verlag New York.

Wu, Chi-Hong, Claudia Fallini, Nicola Ticozzi, Pamela J. Keagle, Peter C. Sapp, Katarzyna Piotrowska, Patrick Lowe, et al. 2012. “Mutations in the Profilin 1 Gene Cause Familial Amyotrophic Lateral Sclerosis.” Nature 488 (7412): 499–503.

Yamanaka, Koji, Seung Joo Chun, Severine Boillee, Noriko Fujimori-Tonou, Hirofumi Yamashita, David H. Gutmann, Ryosuke Takahashi, Hidemi Misawa, and Don W. Cleveland. 2008. “Astrocytes as Determinants of Disease Progression in Inherited Amyotrophic Lateral Sclerosis.” Nature Neuroscience 11 (3): 251–53.

Yang, Jian, Beben Benyamin, Brian P. McEvoy, Scott Gordon, Anjali K. Henders, Dale R. Nyholt, Pamela A. Madden, et al. 2010. “Common SNPs Explain a Large Proportion of the Heritability for Human Height.” Nature Genetics 42 (7): 565–69.

Yin, Tengfei, Dianne Cook, and Michael Lawrence. 2012. “Ggbio: An R Package for Extending the Grammar of Graphics for Genomic Data.” Genome Biology 13 (8): R77.

Young, Adam M. H., Natsuhiko Kumasaka, Fiona Calvert, Timothy R. Hammond, Andrew Knights, Nikolaos Panousis, Jun Sung Park, et al. 2021. “A Map of Transcriptional Heterogeneity and Regulatory Variation in Human Microglia.” Nature Genetics 53 (6): 861–68.

Yu, Guangchuang, Li-Gen Wang, Yanyan Han, and Qing-Yu He. 2012. “clusterProfiler: An R Package for Comparing Biological Themes Among Gene Clusters.” OMICS: A Journal of Integrative Biology. https://doi.org/10.1089/omi.2011.0118.

Zamanian, Jennifer L., Lijun Xu, Lynette C. Foo, Navid Nouri, Lu Zhou, Rona G. Giffard, and Ben A. Barres. 2012. “Genomic Analysis of Reactive Astrogliosis.” The Journal of Neuroscience: The Official Journal of the Society for Neuroscience 32 (18): 6391–6410.

Zeng, Biao, Jaroslav Bendl, Roman Kosoy, John F. Fullard, Gabriel E. Hoffman, and Panos Roussos. 2021. “Trans-Ethnic eQTL Meta-Analysis of Human Brain Reveals Regulatory Architecture and Candidate Causal Variants for Brain-Related Traits.” medRxiv. https://www.medrxiv.org/content/10.1101/2021.01.25.21250099v1.abstract.

Zhao, Weihua, Wenjie Xie, Weidong Le, David R. Beers, Yi He, Jenny S. Henkel, Ericka P. Simpson, Albert A. Yen, Qin Xiao, and Stanley H. Appel. 2004. “Activated Microglia Initiate Motor Neuron Injury by a Nitric Oxide and Glutamate-Mediated Mechanism.” Journal of Neuropathology and Experimental Neurology 63 (9): 964–77.

Zondler, Lisa, Kathrin Müller, Samira Khalaji, Corinna Bliederhäuser, Wolfgang P. Ruf, Veselin Grozdanov, Meinolf Thiemann, et al. 2016. “Peripheral Monocytes Are Functionally Altered and Invade the CNS in ALS Patients.” Acta Neuropathologica 132 (3): 391–411.

