## Supplementary Figures 1-20 for "Integrative genetic analysis of the amyotrophic lateral sclerosis spinal cord implicates glial activation and suggests new risk genes"

### Table of Contents


Supplementary Figure 1: Quality control of 1,917 RNA-seq samples. 3

Supplementary Figure 2: RNA-seq quality control continued. 4

Supplementary Figure 3: Defining sources of variation in the spinal cord samples. 5

Supplementary Figure 4: Motor neuron marker genes are lowly expressed. 6

Supplementary Figure 5: Correlating differentially expressed genes between each tissue. 7

Supplementary Figure 6: Full pathway enrichment results. 8

Supplementary Figure 7: Activation gene sets partially overlap. 9

Supplementary Figure 8: Full deconvolution plots. 10

Supplementary Figure 9: Deconvolution plots continued. 11

Supplementary Figure 10: Comparing deconvolution estimates between tools. 12

Supplementary Figure 11: Comparing deconvolution estimates between references. 13

Supplementary Figure 12: Expression-Weighted Cell-type Enrichment (EWCE) analysis. 14

Supplementary Figure 13: Comparing C9orf72 ALS to Sporadic ALS. 15

Supplementary Figure 14: Correlating disease duration associations between sections. 16

Supplementary Figure 16: Whole genome sequencing QC. 17

Supplementary Figure 16: QTL discovery rate with different numbers of PEER factors. 18

Supplementary Figure 17: Transcriptome-wide Association Study (TWAS). 19

Supplementary Figure 18: ATXN3 repeat lengths in the NYGC ALS WGS cohort. 20

Supplementary Figure 19: Cell-type fidelity scores for each prioritised gene. 21

Supplementary Figure 20: Cell-type proportion correlation in control samples. 21

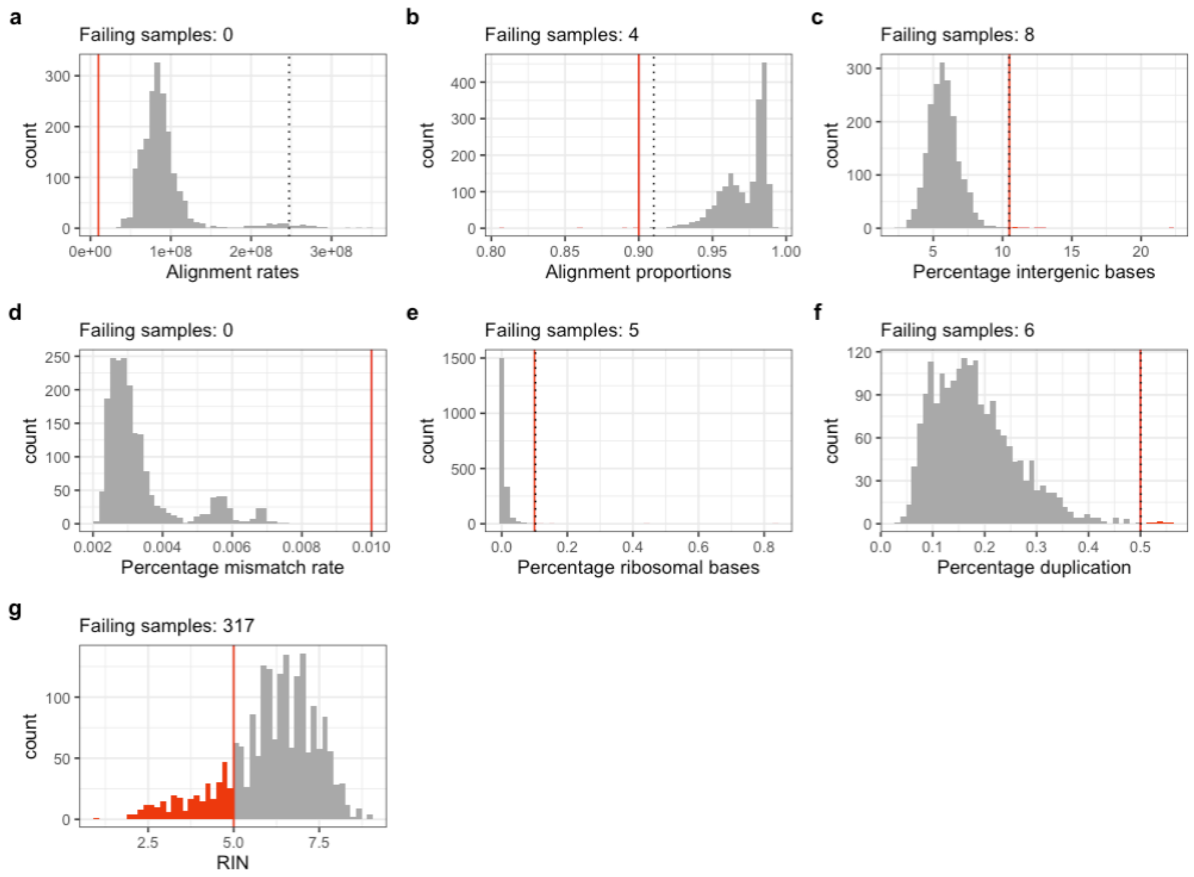


**Supplementary Figure 1: Quality control of 1,917 RNA-seq samples.**
Tthe January 2020 data freeze of the NYGC ALS consortium cohort, comprising post-mortem RNA-seq from cortex, spinal cord and cerebellum. (a-g) Distributions of different metrics of sequencing quality, as ascertained by Picard tools. Dotted lines denote 4 standard deviations from the mean value, and the red line denotes the threshold imposed. Red bars indicate the samples flagged by different thresholds. Note that the RIN > 5 threshold was applied for the differential expression analysis but not the QTL analysis.

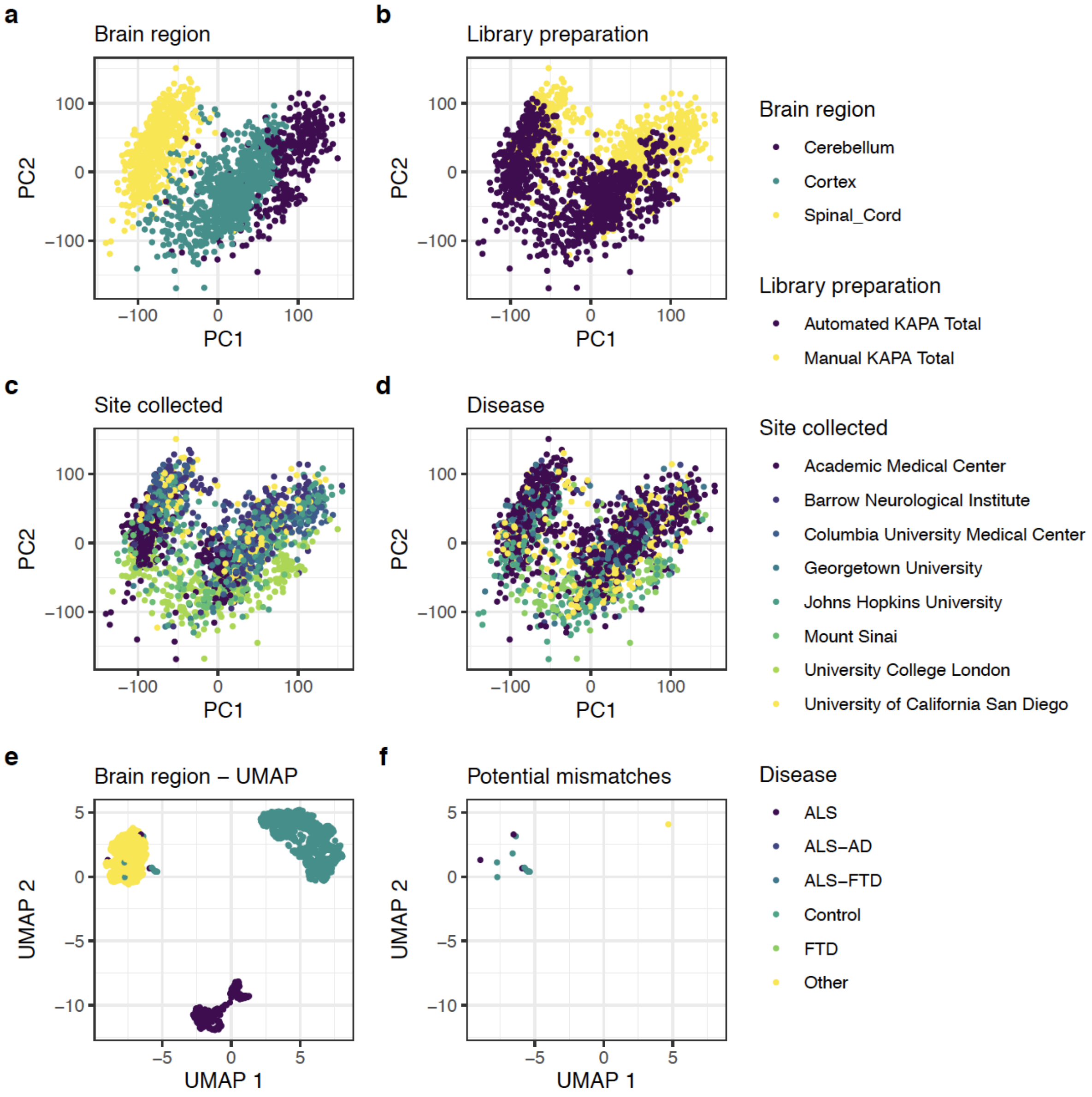


**Supplementary Figure 2: RNA-seq quality control continued.**
(a-d) Principal component analysis of voom-transformed gene expression from 1,792 cerebellum, cortex and spinal cord samples, showing separation by brain region (a), library preparation method (b), submitting site (c) and disease status (d). (e-f) UMAP clustering of same samples, coloured by recorded brain region (e), and with potential tissue swaps highlighted (f). These samples were removed from further analysis. Only the spinal cord samples were used in this study.

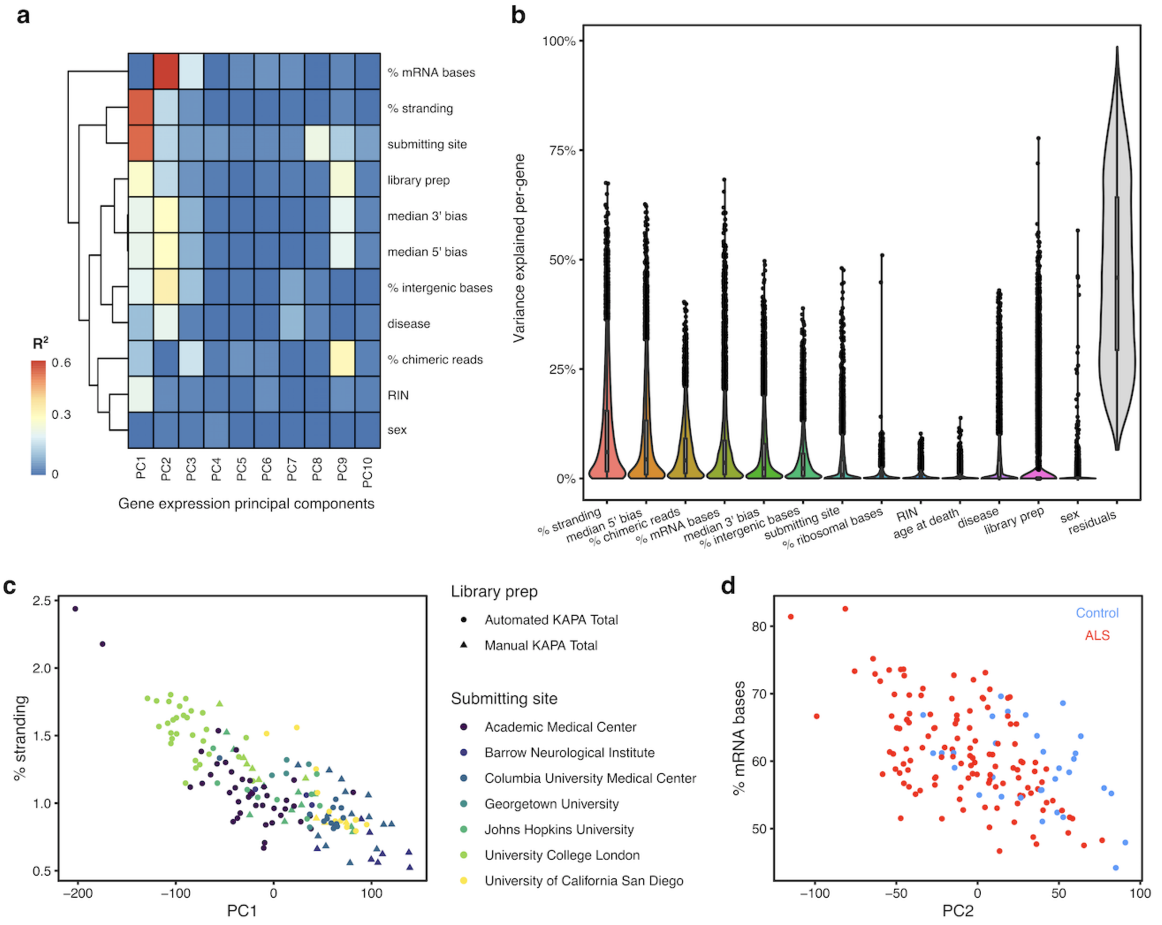


**Supplementary Figure 3: Defining sources of variation in the spinal cord samples.** Lumbar spinal cord samples shown for illustration. a) Comparing the top 10 principal components (PC) of gene expression with technical and clinical variables using linear regression. $R^2$ from univariate linear regression of each PC against each variable. The first two principal components are strongly explained by technical factors and submitting sites. The top 5 genotyping PCs showed no association with any expression PC and so are omitted from this figure. b) VariancePartition computes variance explained per gene for each variable independently. Boxplots show the median, first and third quartile of the distribution. Jittered points are genes outside 1.5 times the interquartile range. More variance is explained on average by technical factors than clinical factors. (c-d) Example associations. c) PC1 correlates with stranding percentage, the proportion of the total transcript-mapping reads being from the wrong strand. This is likely caused by a change in library preparation method which coincided with the sequencing of samples from different sites. d) PC2 correlates with both % mRNA bases, the proportion of sequenced bases aligned to mRNA transcripts, and case-control status.

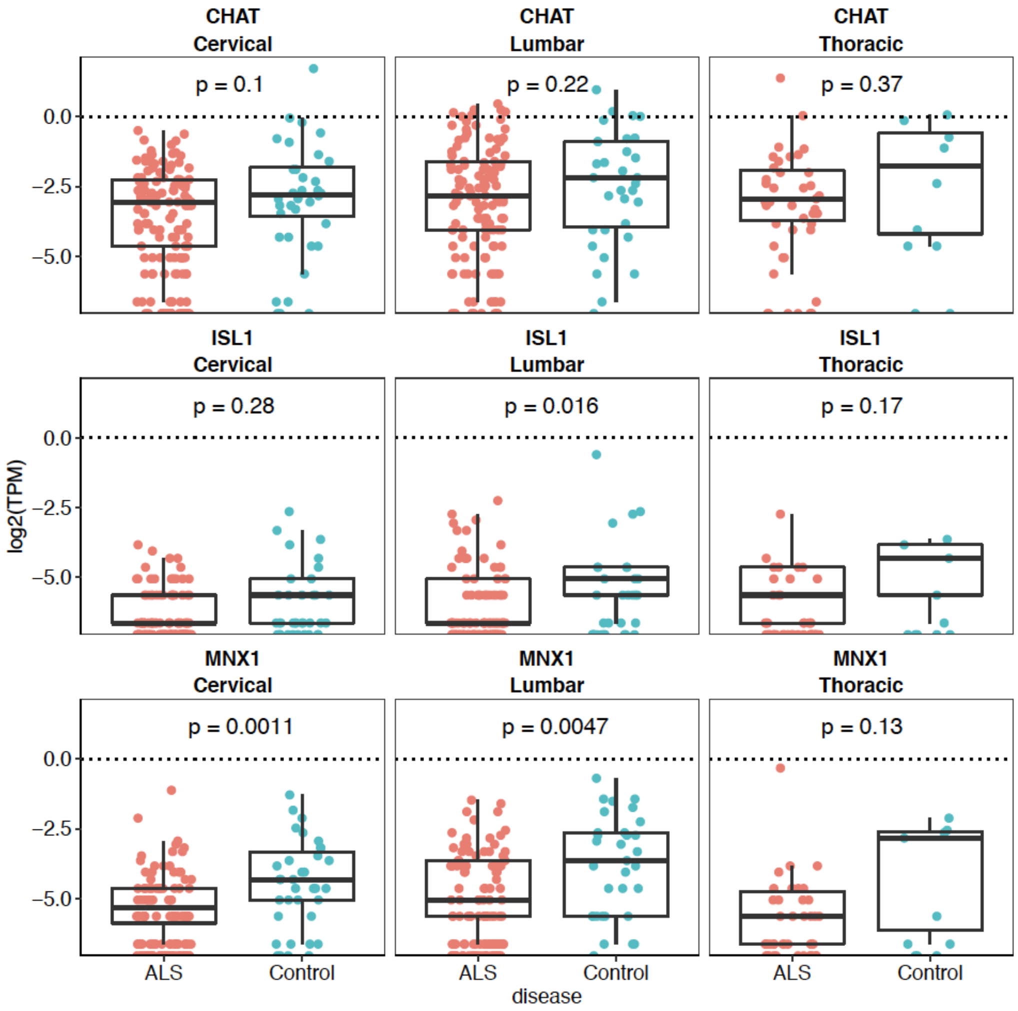


**Supplementary Figure 4: Motor neuron marker genes are lowly expressed.** Expression, in $\log_2$(transcripts per million) for motor neuron marker genes CHAT, ISL1 and MNX1 in the spinal cords of ALS and Control samples. The dotted line is TPM=1. P-values are from uncorrected wilcoxon non-parametric tests.

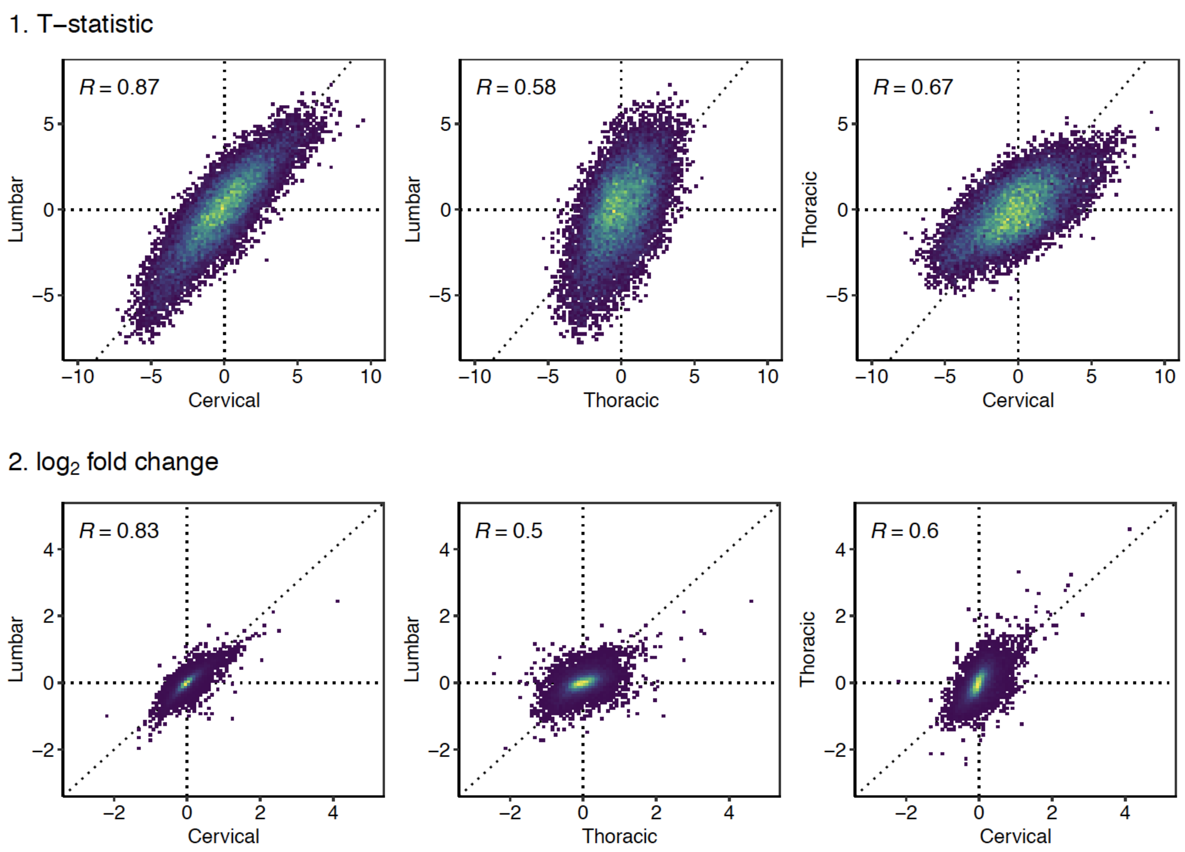


**Supplementary Figure 5: Correlating differentially expressed genes between each tissue.**
Upper plot: T-statistics, representing the sign and magnitude of the differential expression test for each gene compared between each pair of tissues. Pearson correlation used. Lower plot: $\log_2$ fold changes, representing the estimated effect size of the difference in expression between ALS samples and controls. Individual points are genes, colour refers to density of overlapping points.

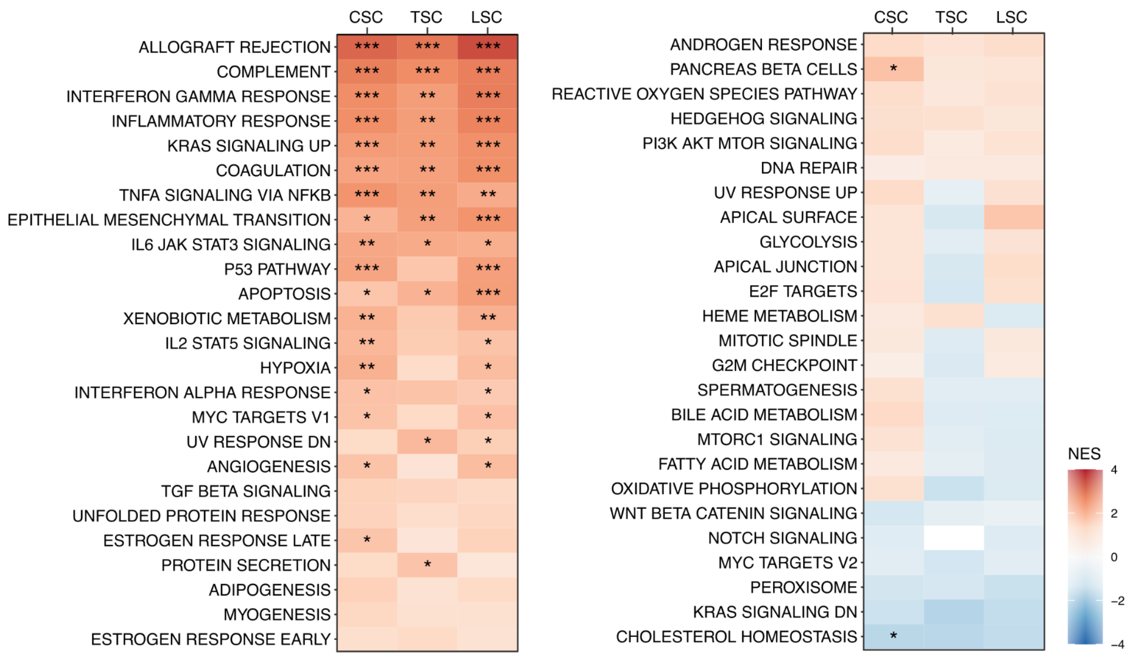


**Supplementary Figure 6: Full pathway enrichment results.**
Gene set enrichment analysis results for all molecular signatures database (MSigDb) hallmark pathways in the three spinal cord sections. Tiles are coloured by the normalised enrichment score (NES). *** q < 1e-4; ** q < 1e-3; * q < 0.05; . q > 0.05.

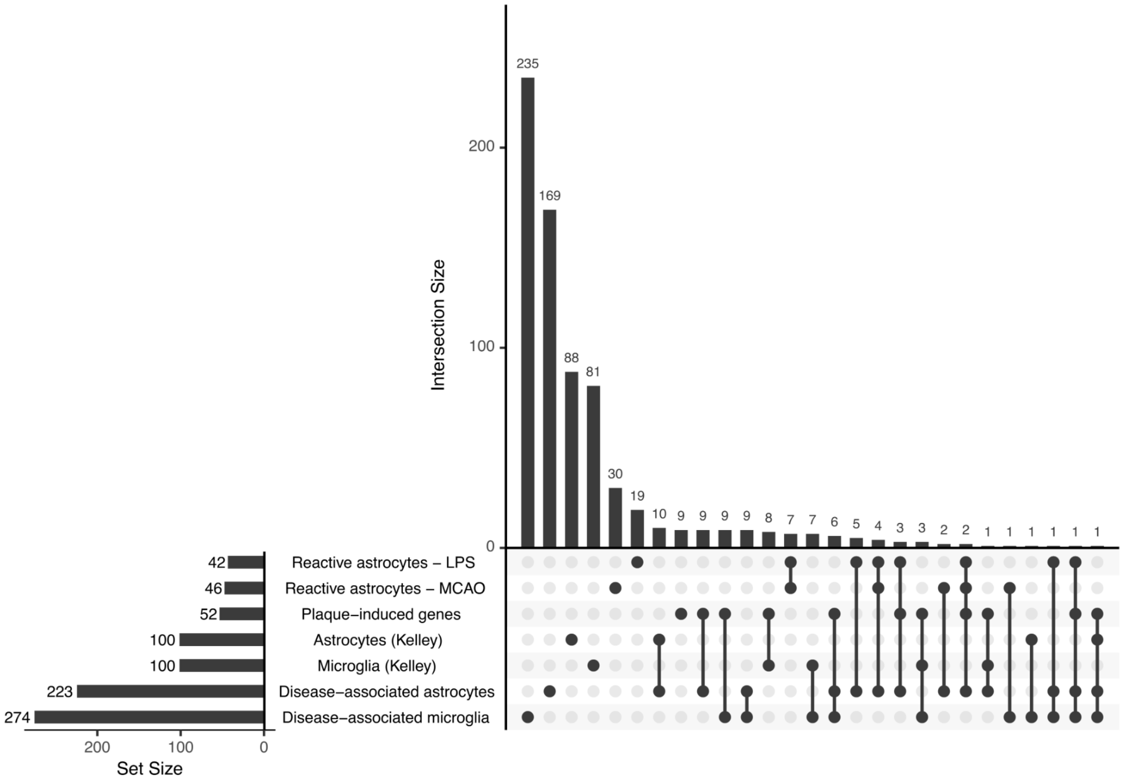


**Supplementary Figure 7: Activation and glia gene sets partially overlap.**
Upset plot to compare number of genes in each set (horizontal bars) with the intersections between each set (vertical bars). Non-overlapping intersections labelled as filled circles. Intersections indicated by lines connecting circles. Sets are largely distinct, with the exception of plaque-induced genes (9 of 52 genes are unique to the set).

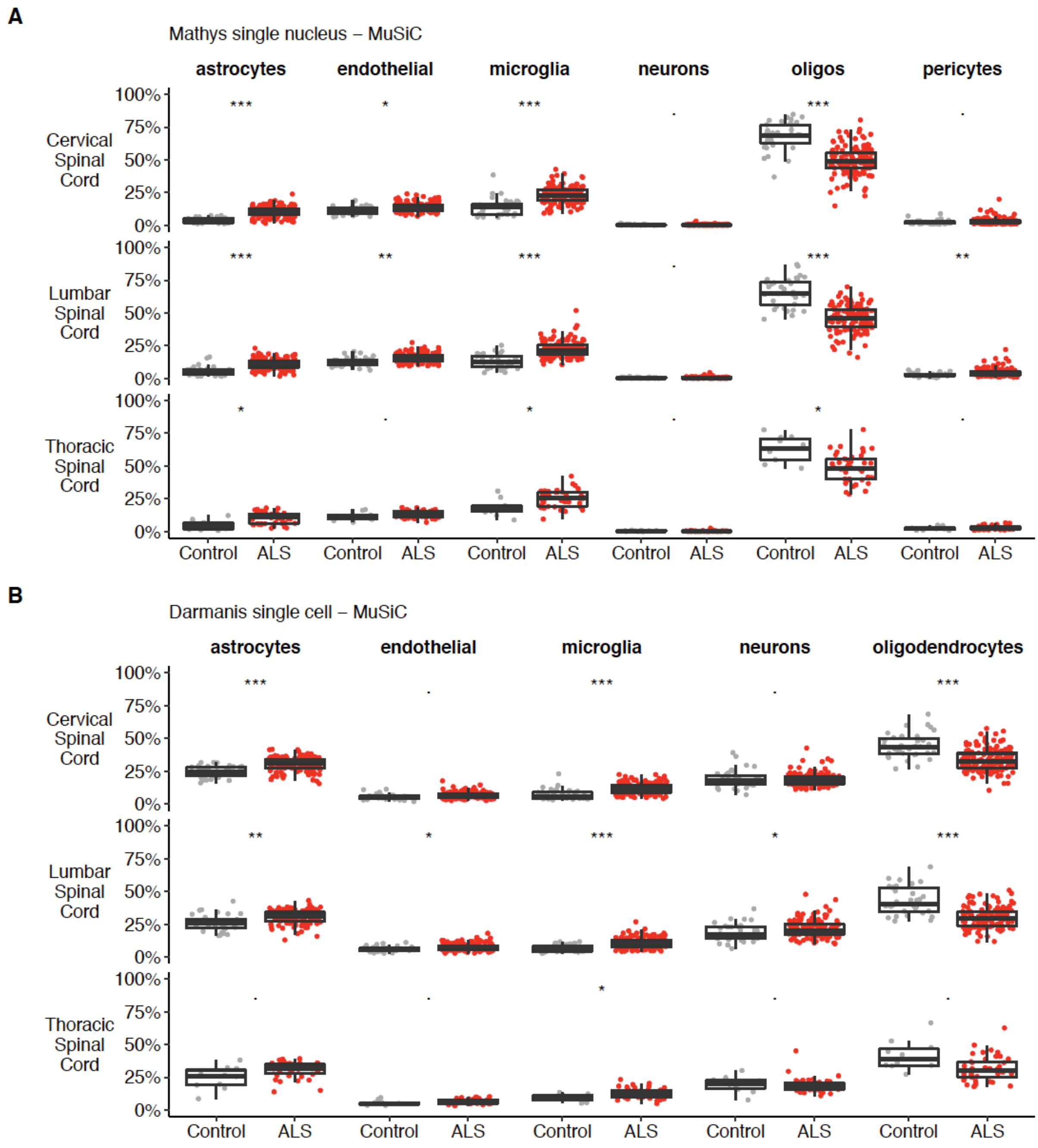


**Supplementary Figure 8: Full deconvolution plots.**
a: Cell-type proportion estimates in each sample from MuSiC, using the Mathys et al single nucleus RNA-seq data. Excitatory neurons only plotted. b: Estimates from MuSiC using the Darmanis et al single cell RNA-seq. Asterisks refer to Bonferroni-corrected P-values, q, from a Wilcoxon test comparing each cell-type estimate between ALS and Control, residualized after regressing out technical covariates. *** q < 1e-4; ** q < 1e-3; * q < 0.05; . q > 0.05.

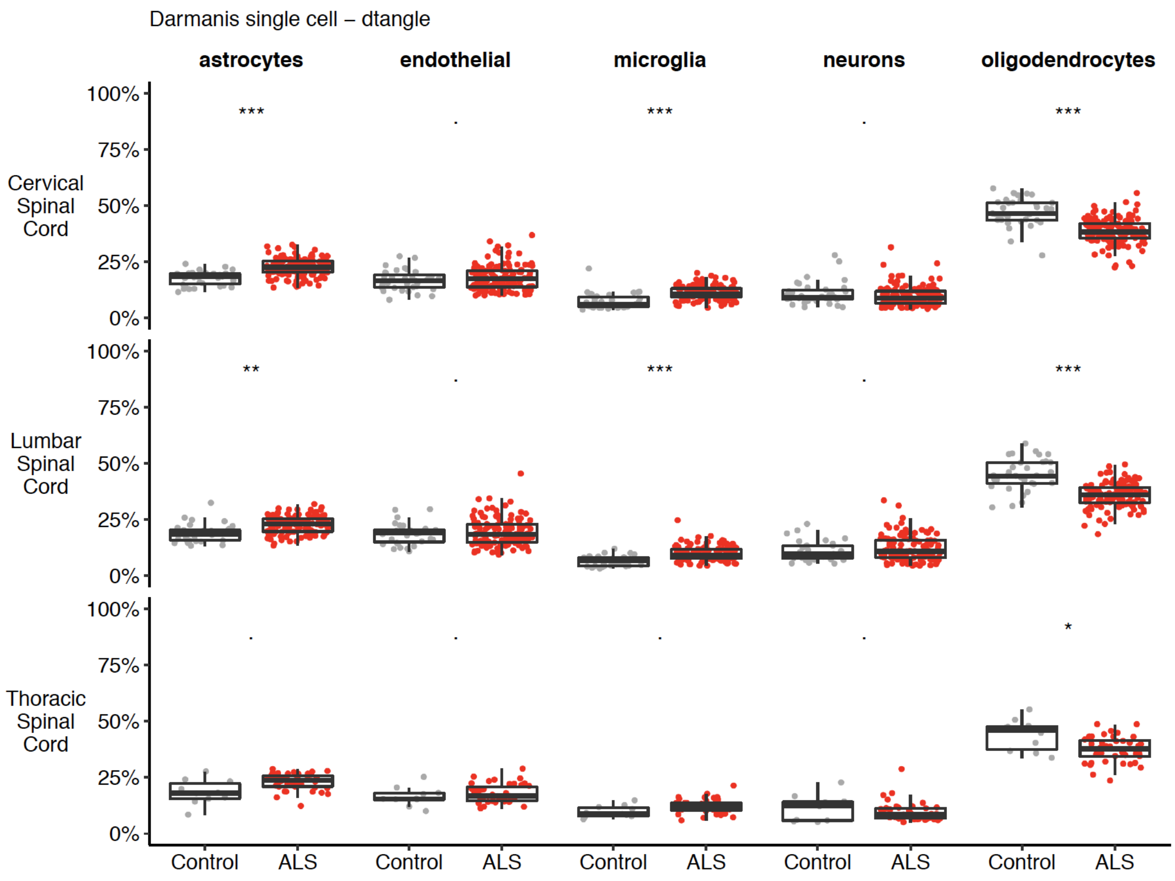


**Supplementary Figure 9: Deconvolution plots continued.**
Cell-type proportion estimates in each sample from dtangle using the Darmanis et al single cell RNA-seq. Asterisks refer to Bonferroni-corrected P-values, q, from a Wilcoxon test comparing each cell-type estimate between ALS and Control, residualized after regressing out technical covariates. *** $q < 1e-4$; ** $q < 1e-3$; * $q < 0.05$; . $q > 0.05$.

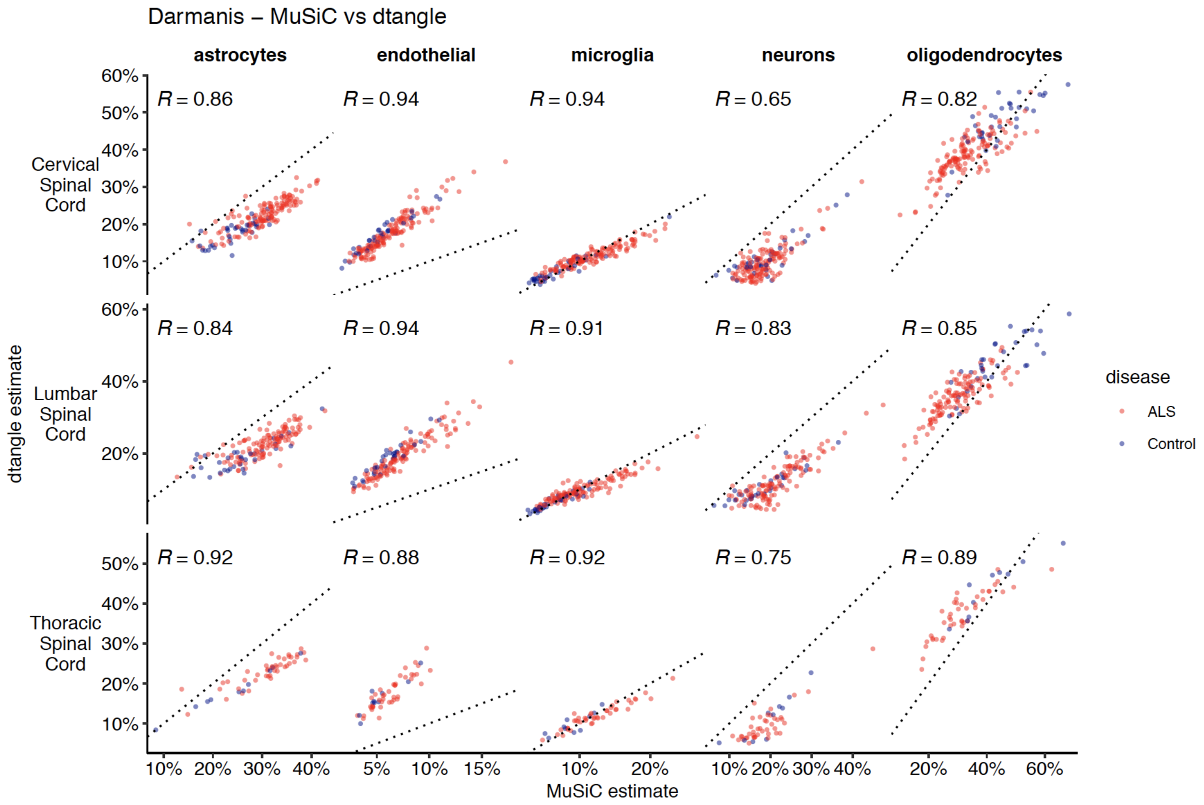


**Supplementary Figure 10: Comparing deconvolution estimates between tools.** Comparing deconvolution estimates between the MuSiC and dtangle algorithms using the same Darmanis et al single cell RNA-seq reference data. Dotted line is x=y, R refers to Spearman correlation. dtangle was not run with the Mathys dataset.

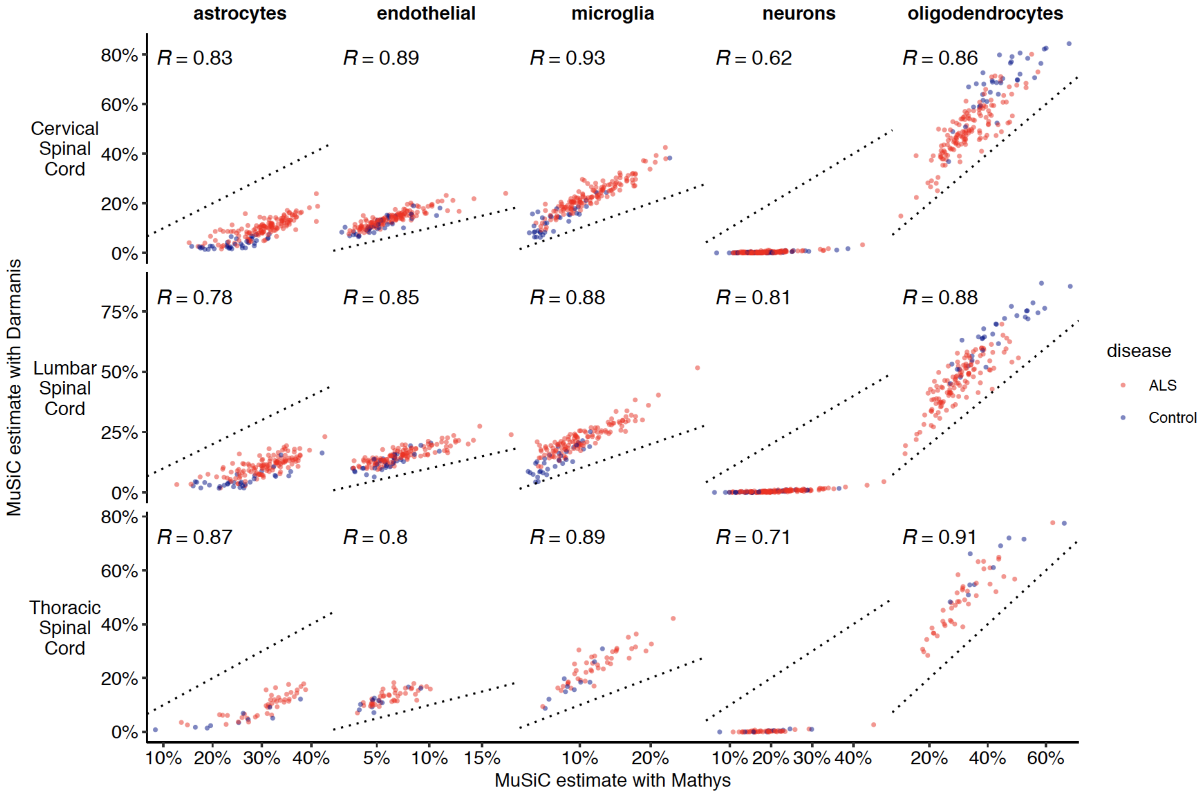


**Supplementary Figure 11: Comparing deconvolution estimates between references.** Comparing deconvolution estimates from MuSiC between the Darmanis and Mathys reference panels. Dotted line is x=y, R refers to Spearman correlation.

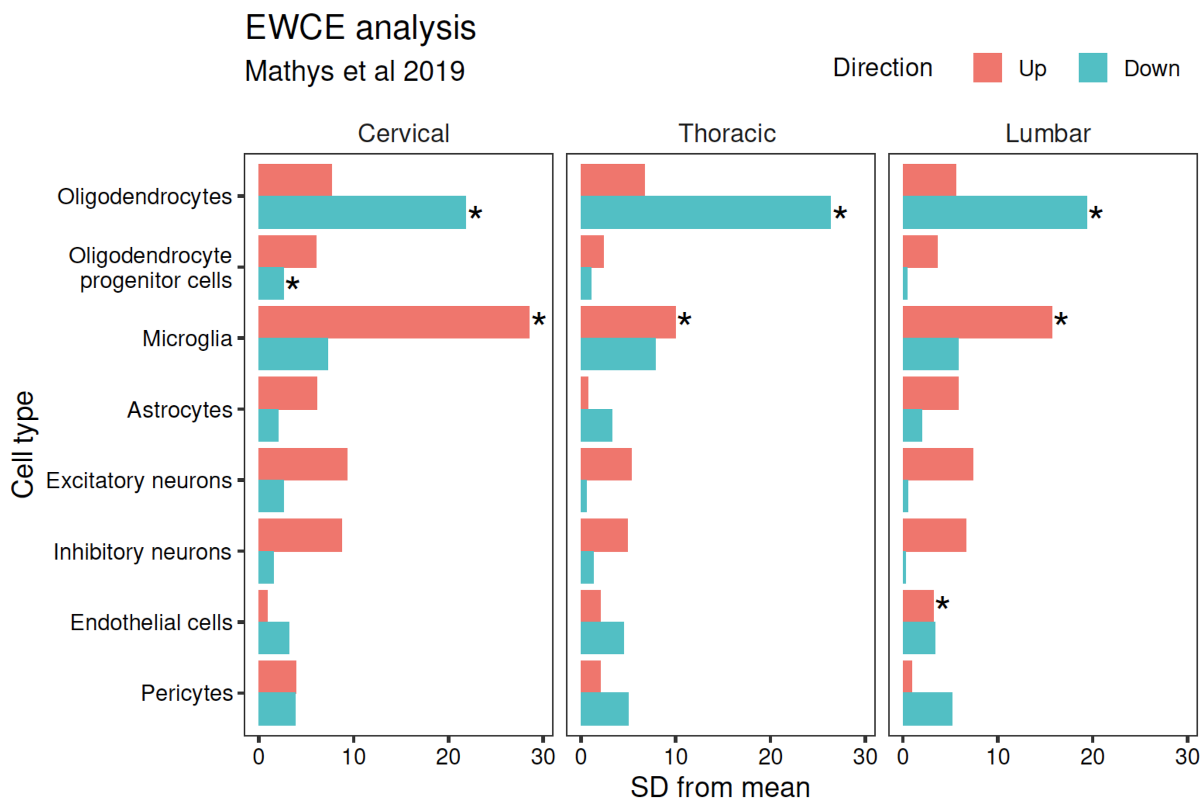


**Supplementary Figure 12: Expression-Weighted Cell-type Enrichment (EWCE) analysis.**
Results of EWCE analysis ran using the top 250 up- and down-regulated differentially expressed genes in each tissue and a panel of gene cell-type specificity scores created from single-nucleus RNA-seq from the human cortex. Results presented as numbers of standard deviations (SD) from the mean of the empirical null distribution from 10,000 permuted gene sets. Enrichments within upregulated genes in red, downregulated genes in blue.

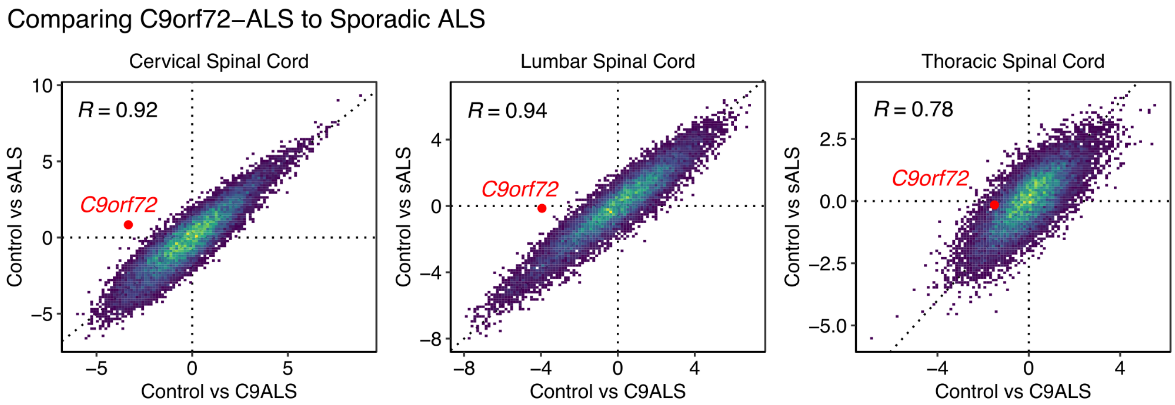


**Supplementary Figure 13: Comparing C9orf72 ALS to Sporadic ALS.**
T-statistics, representing the sign and magnitude of the differential expression test for each gene, compared between an analysis of C9orf72-ALS vs Controls (x-axis) to an analysis of Sporadic ALS vs Controls (y-axis). Pearson correlation used. *C9orf72* gene highlighted in red. Individual points are genes, colour refers to density of overlapping points.

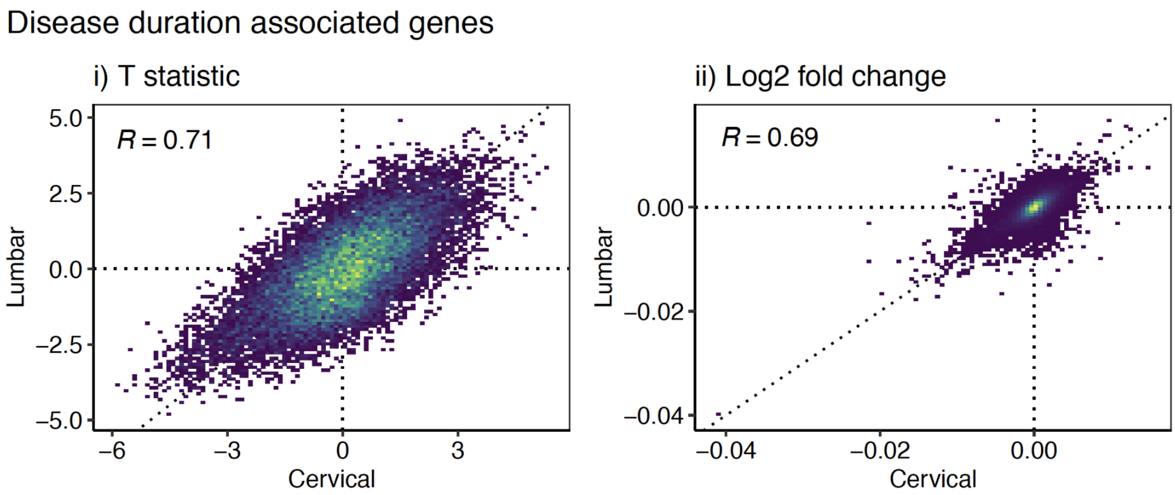


**Supplementary Figure 14: Correlating disease duration associations between sections.**

i) Using the test statistics for each gene. ii) Using the $\log_2$ fold changes for each gene. R refers to Pearson correlation coefficients.

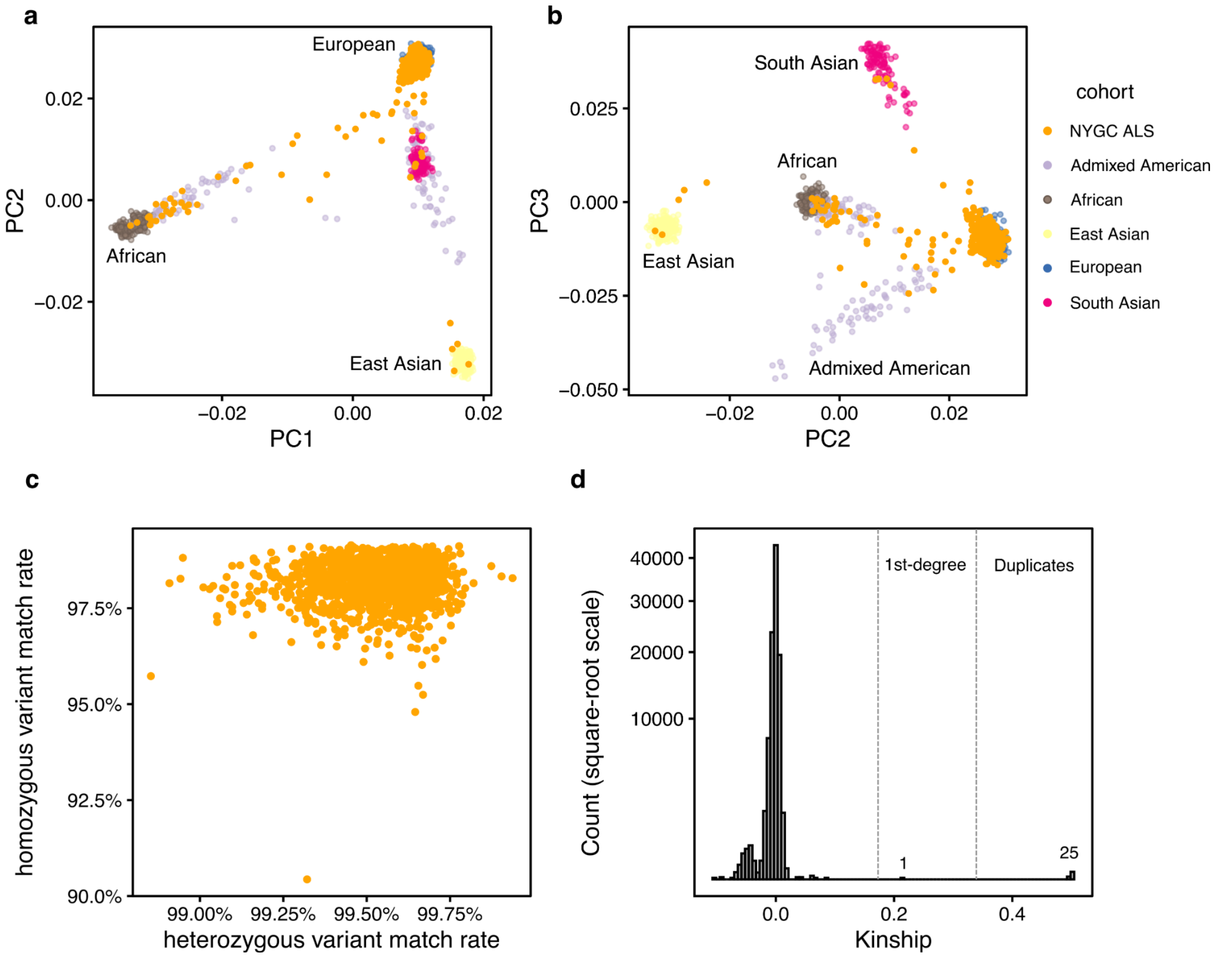


**Supplementary Figure 15: Whole genome sequencing QC.**
a-b) Genotype principal component analysis of NYGC ALS consortium donors projected on to 1000 Genomes phase 3 superpopulations. Majority of donors cluster with European superpopulation. c) All donors match the correct RNA-seq sample by genotype, from MBV. d) Kinship estimates from KING. Kinship > 0.34 indicates monozygotic twins or duplicates. 25 pairs of duplicate donors and 1 first degree relative pair identified.

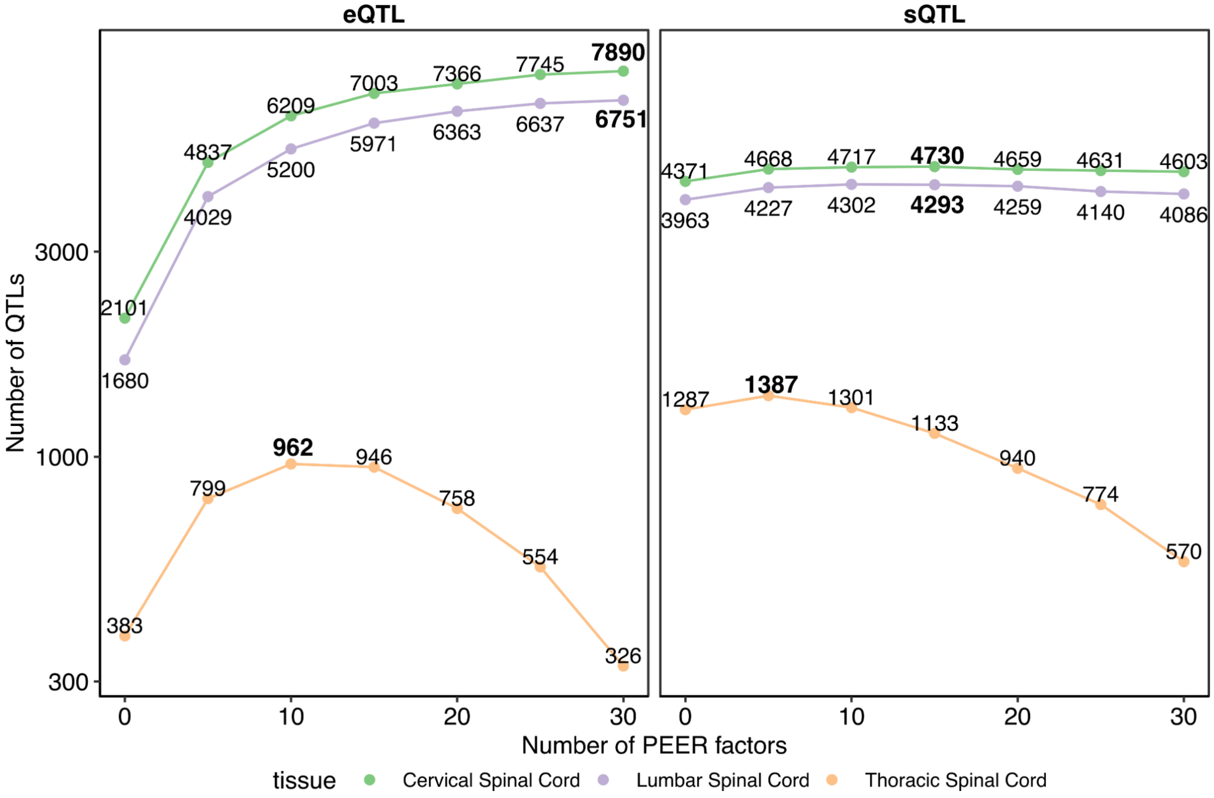


**Supplementary Figure 16: QTL discovery rate with different numbers of PEER factors.** Left panel - number of genes with an eQTL at qvalue < 0.05 in each tissue and PEER threshold. Right panel - number of genes with a splicing QTL at each threshold.

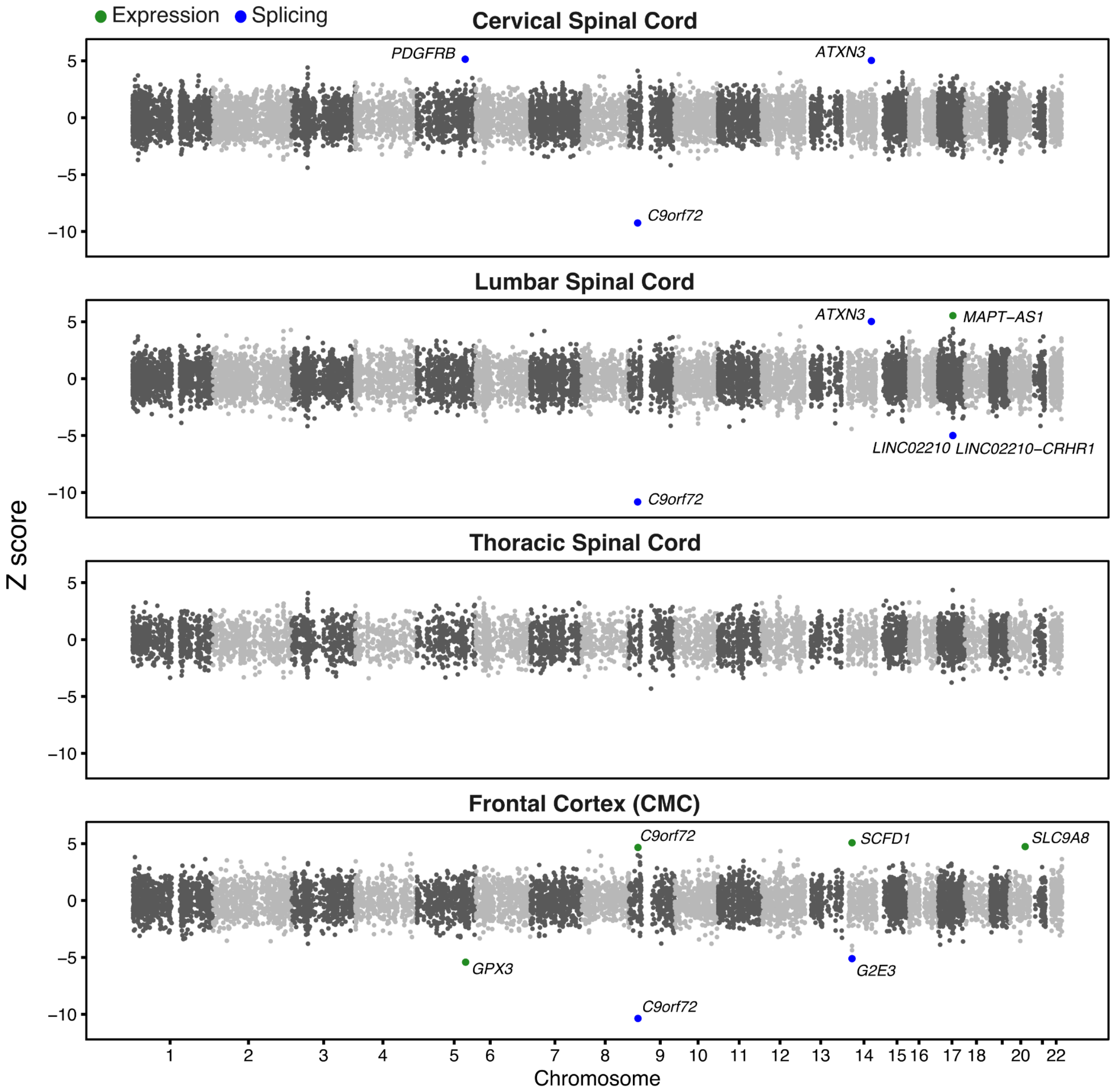


**Supplementary Figure 17: Transcriptome-wide Association Study (TWAS).**
Results from running FUSION TWAS with the Nicolas et al. 2018 ALS GWAS and reference panels computed from the three spinal cord sections, as well as pre-computed weights from the CommonMind Consortium (CMC) dorsolateral prefrontal cortex samples. Each dot is a gene. Genes with an FDR < 0.05 are labelled and coloured by whether they came from an expression (green) or splicing (blue) model. For the splicing models, as multiple splice junction models were created per gene, the model with the largest absolute Z-score is plotted.

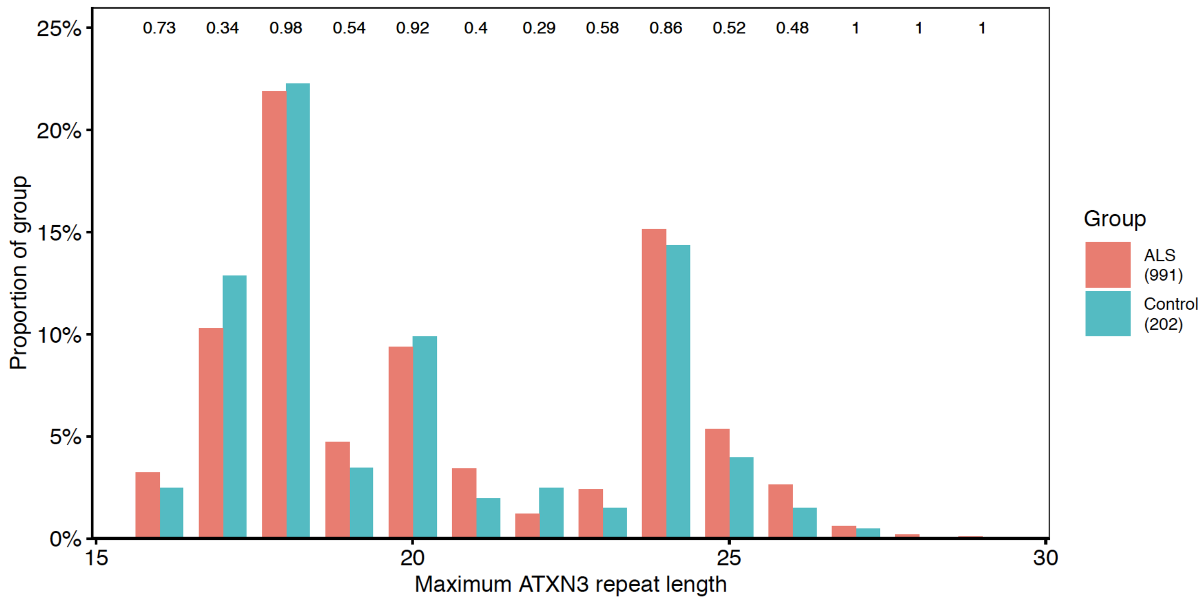


**Supplementary Figure 18: ATXN3 repeat lengths in the NYGC ALS WGS cohort.** Repeat lengths measured with ExpansionHunter. For each individual the maximum repeat length was taken and plotted as a proportion of the total of either cases or controls. Each repeat length was compared with a Chi-squared test of equal proportions. Numbers at top of plot are nominal P-values.

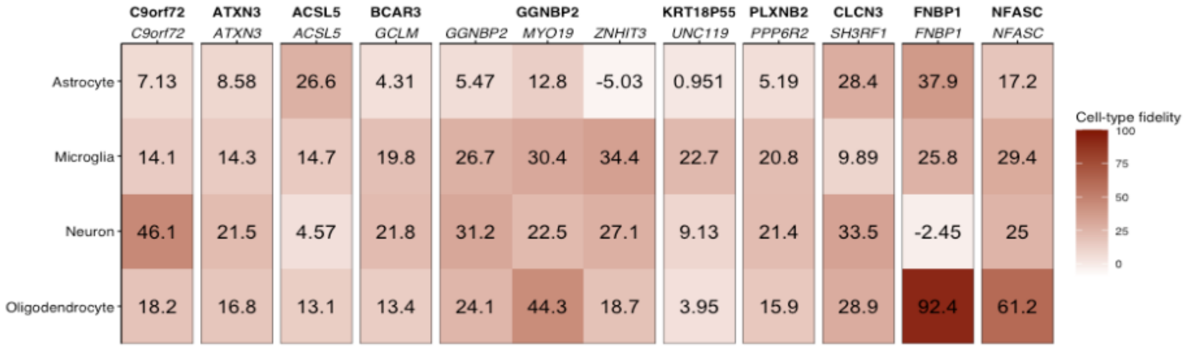


**Supplementary Figure 19: Cell-type fidelity scores for each prioritised gene.** Scores from Kelley et al. (see methods).

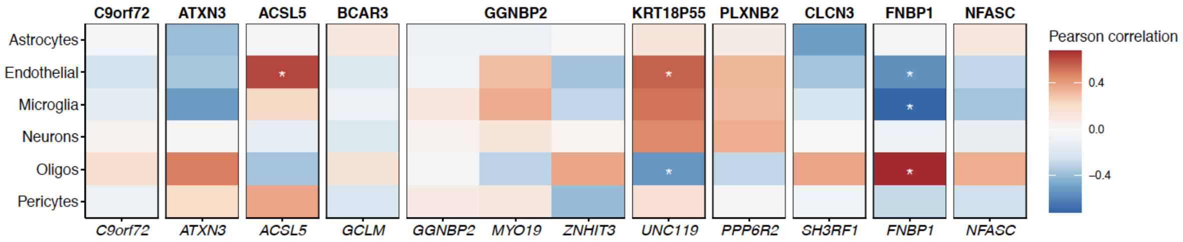


**Supplementary Figure 20: Cell-type proportion correlation in control cervical spinal cord samples.**

Voom-normalised gene expression values in control samples only correlated with estimated cell-type proportions (see methods).
